# A novel alpha-synuclein G14R missense variant is associated with atypical neuropathological features

**DOI:** 10.1101/2024.09.23.24313864

**Authors:** Christof Brücke, Mohammed Al-Azzani, Nagendran Ramalingam, Maria Ramón, Rita L. Sousa, Fiamma Buratti, Michael Zech, Kevin Sicking, Leslie Amaral, Ellen Gelpi, Aswathy Chandran, Aishwarya Agarwal, Susana R. Chaves, Claudio O. Fernández, Ulf Dettmer, Janin Lautenschläger, Markus Zweckstetter, Ruben Fernandez Busnadiego, Alexander Zimprich, Tiago Fleming Outeiro

## Abstract

**Background:** Parkinson’s disease (PD) affects millions of people worldwide, but only 5–10% of patients suffer from a monogenic form of the disease with Mendelian inheritance. *SNCA,* the gene encoding for the protein alpha-synuclein (aSyn), was the first to be associated with familial forms of PD and, since then, several missense variants and multiplications of the *SNCA* gene have been established as rare causes of autosomal dominant forms of PD.

**Aim and methods:** A patient carrying aSyn missense mutation and his family members were studied. We present the clinical features, genetic testing - whole exome sequencing (WES), and neuropathological findings. The functional consequences of this aSyn variant were extensively investigated using biochemical, biophysical, and cellular assays.

**Results:** The patient exhibited a complex neurodegenerative disease that included generalized myocloni, bradykinesia, dystonia of the left arm and apraxia. WES identified a novel heterozygous *SNCA* variant (cDNA 40G>A; protein G14R). Neuropathological examination showed extensive atypical aSyn pathology with frontotemporal lobar degeneration (FTLD) and nigral degeneration pattern with abundant ring-like neuronal inclusions, and few oligodendroglial inclusions. Sanger sequencing confirmed the *SNCA* variant in the healthy, elderly parent of the patient patient suggesting incomplete penetrance. NMR studies suggest that the G14R mutation induces a local structural alteration in aSyn, and lower thioflavin T binding in in vitro fibrillization assays. Interestingly, the G14R aSyn fibers display different fibrillar morphologies as revealed by cryo-electron microscopy. Cellular studies of the G14R variant revealed increased inclusion formation, enhanced membrane association, and impaired dynamic reversibility of serine-129 phosphorylation.

**Summary:** The atypical neuropathological features observed, which are reminiscent of those observed for the G51D aSyn variant, suggest a causal role of the *SNCA* variant with a distinct clinical and pathological phenotype, which is further supported by the properties of the mutant aSyn, compatible with the strain hypothesis of proteinopathies.

## Introduction

Parkinsońs disease (PD) is a common neurodegenerative disease that leads to motor symptoms of bradykinesia, tremor and rigidity and concomitant non-motor symptoms.^1^ Around 5-10% of patients suffer from a monogenetic form of PD. The neuropathological hallmarks of PD are the loss of dopaminergic neurons in the substantia nigra and the accumulation of intraneuronal inclusions known as Lewy bodies (LBs) that are enriched in the protein alpha-synuclein (aSyn).^2–5^ aSyn also accumulates in protein inclusions in other neurodegenerative diseases, such as dementia with Lewy bodies (DLB) and multiple system atrophy (MSA). However, it is now becoming evident that the structural arrangement of aSyn in inclusions differs depending on the synucleinopathy.^6–8^

aSyn, encoded by the *SNCA* gene, is an intrinsically disordered protein abundant in the brain, in red blood cells, and in several other cell types, but its precise function is still unclear. In neurons, aSyn localizes mainly in the pre-synaptic compartment, but can also be found in cytoplasm, the nucleus, mitochondria, endoplasmic reticulum and associated with membranes.^9,10^

Abnormal folding, accumulation, and aggregation of aSyn perturbs cellular homeostasis, leading to cellular pathologies thought to culminate in cell death, especially in dopaminergic neurons in the substantia nigra, for reasons we do not fully understand. While aSyn aggregation has been traditionally seen as the culprit leading to neuronal dysfunction and death, it is also possible that loss of aSyn function, known as proteinopenia, contributes to disease.^11,12^

Several variants of aSyn have been linked to familial forms of PD and include, for example, A53T, V15A, A30G, A30P, E46K, H50Q, G51D, A53E, A53V and T72M, highlighting the importance of aSyn in PD.^2,5,13–20^ Strikingly, the aSyn-related phenotypic spectrum appears to be broader than initially described, with marked differences reported for different variants.^21,22^ For example, some of the aSyn variants seem to lead to early onset PD with a higher probability of developing early cognitive deficits. In some cases, the penetrance of the gene variants seems to be incomplete, and the pathogenicity of some aSyn variants is still controversial.^19,21,23^ Interestingly, although most of the aSyn mutations linked to PD reside in the N-terminal region, a local change in aSyn structure near a single mutation site can have a profound effect on its aggregation as well as on its physiological properties, including phosphorylation at S129 (pS129).^24,25^ *In vitro* studies have shown divergent aggregation properties for these mutations, with enhanced aggregation propensity for some, such as E46K, and attenuated propensity for others, such as A30P.^26–28^ Therefore, although aSyn missense mutations are rare, studying the effects of the various mutations on aSyn greatly contributes to our understanding of both aSyn biology and pathobiology in PD and other synucleinopathies.

Here, we report the identification of a new heterozygous *SNCA* variant (cDNA40G>A; protein G14R) in an Austrian family, and describe clinical features, genetic findings, functional effects on aSyn, and the neuropathology of a deceased patient.

## Materials and methods

We evaluated the patient at the Department of Neurology of the Medical University of Vienna, Austria, in a study with approval of the ethics committee of the Medical University of Vienna (EK1844/2019), and with written informed consent to participate in the genetic research described in this study by all subjects involved. Comprehensive general and neurological examinations were conducted.

### Genetic testing

Due to the atypical phenotype genetic testing was ordered. Informed consent was obtained from patients and members of the family. Genomic DNA was isolated from peripheral blood using a standard protocol. Sequencing was performed at the Institute of Human Genetics of the Technical University of Munich, Germany. The patient’s sample was enriched using Sure Select Human All Exon Kit (Agilent 60mb V6) and sequencing was carried out on an Illumina NovaSeq6000 system (Illumina, San Diego, California). The average exome coverage was 122x; 98% of the target regions were covered at least 20x and 100% of the SNCA region was covered with at least 25x. Sanger sequencing was used to confirm the G14R mutation.

### Neuropathology

Neuropathological study was performed according to standard procedures at the Division of Neuropathology and Neurochemistry. Full details of the experimental approach are included in the supplementary materials.

### Biophysical and cellular studies

In order to check functional consequences of the G14R mutation on aSyn structural and aggregational properties, we performed Nuclear magnetic resonance (NMR) spectroscopy, in vitro thioflavin T (ThT) fluorescence-based aggregation assays, and cryo-EM imaging and analysis of WT and G14R fibrils using recombinantly produced human aSyn. In addition, we tested in cellular models how G14R mutation could alter its aggregation tendency, condensate formation, membrane binding, and S129 photoporation status. Methods for the different assays are described in the supplementary material file.

## Results

### Clinical presentation with atypical features

The index patient was studied and treated at the neurological outpatient clinic and family members (both parents, two siblings and a child) were examined and genetically tested. At the age of 50s, the patient developed a stuttering speech with palilalia, myoclonic jerks and action tremor of the limbs and an abnormal gait. Due to the fluctuating presentation and worsening during emotional stress, a functional disorder was initially suspected. On follow-up, the patient showed severe bradykinesia, dystonia of the left arm and later a limb apraxia. L-dopa treatment had only a slight and temporary effect on bradykinesia and was, therefore, stopped after a few months. Screaming, laughing, and kicking with the legs during sleep were reported by the spouse, and a REM sleep behavior disorder (RBD) was suspected. A diagnostic sleep study (polysomnography) was not performed since the patient was too disabled at this time point. Clonazepam improved the RBD like-symptoms. Symptoms progressed and the patient developed a severe apathy. Five MRI scans of the patient, performed 3 and 4 years after disease onset, were unremarkable. Ioflupane (FP-CIT, DaTScan) single-photon emission computed tomography (SPECT) performed around 4 years after first symptoms showed a marked bilateral reduction of dopamine transporter activity. FDG-positron emission tomography (PET) showed a bilateral frontal, postcentral, precuneal and basal ganglia (right side) hypometabolism. A tilt-table-test showed no evidence of autonomic dysfunction. Neuropsychological tests initially showed no abnormalities, on follow-up a slowing of information processing and reduced attention were found. CSF showed 12 cells/µl and total protein of 42 mg/dL. After 4 years, the patient lost ambulation and died 2 years later due to complications of pneumonia, after a total disease duration of 6 years.

### Whole-exome sequencing identifies aSyn p.G14R point mutation

To investigate whether a genetic cause was responsible for the patient’s phenotype, we performed whole-exome sequencing (WES). We identified a novel heterozygous variant in the *SNCA* gene (NM_000345.4, cDNA 40G>A; protein G14R) (Figure S1A). We did not find the variant in any publicly accessible database, including GnomAD and the PD Variant Browser.^29^ The G14R aSyn variant is located in an evolutionarily conserved region (Figure S1B) and is predicted as deleterious by all in-silico prediction algorithms (CADD:34, PolyPhen-2: probably damaging, EVE: pathogenic, and alphamissense: pathogenic 0,9832).

The family members were examined for neurological signs by a neurologist trained in movement disorders. Two healthy siblings and a parent tested negative for the variant. The other parent carries the variant (Figure S1A), but was healthy and showed no slowing of movements or non-motor signs hinting at subclinical Lewy body disease (e.g. no RBD, no hyposmia or constipation).

### Neuropathological assessment reveals cortical degeneration and aSyn ring-like structures

After the patient died, an autopsy restricted to the brain was performed. Total brain weight (fixed) was 1300 g. Gross examination showed a fronto-temporal lobar degeneration pattern with severe involvement of the cingulate gyrus and asymmetrical pallor of the substantia nigra. Histology confirmed the FTLD and nigral degeneration with superficial laminar spongiosis in cortical regions, neuronal loss, astrogliosis and microglial reaction associated with an extensive atypical aSyn pathology, i.e. not following classical PD/DLB or MSA cytomorphologies or distribution patterns (Figure 1). The pathology was characterized by abundant ring-like, some comma-shaped and only few spherical neuronal inclusions in superficial and deep cortical layers (frontal-, motor-, and temporal cortex, cingulate, insula, claustrum, basal ganglia (putamen > caudate > pallidum), amygdala, mild involvement of the parietal and nearly no involvement of the occipital cortex and thalamic nuclei) associated with abundant thin and tortuous neurites. Ring-like inclusions were also identified in the granular neurons of the dentate gyrus and ring- and tangle-like inclusions in pyramidal cells mainly of the CA1 sector and parahippocampal region (Figure 1, top panel). Abundant spherical and ring-like inclusions were also detected in the olfactory bulb. The brainstem was comparatively less and only mildly involved but showed also rounded compact and diffuse granular neuronal inclusions in the substantia nigra (SN), periaquaeductal grey, locus coeruleus and only very few and irregular in shape in the reticular formation and raphe, without obvious involvement of the dorsal motor nucleus of the vagal nerve. No typical LBs were observed on HE-stained sections in pigmented nuclei, and the appearance of the inclusions observed was more reminiscent of “pale bodies” (Figure 1, bottom panel). There were also some aSyn positive glial abnormalities reminiscent of MSA glial cytoplasmic inclusions (GCI) in the brainstem white matter, particularly the midbrain. The cerebellum was not affected.

**Figure 1.**
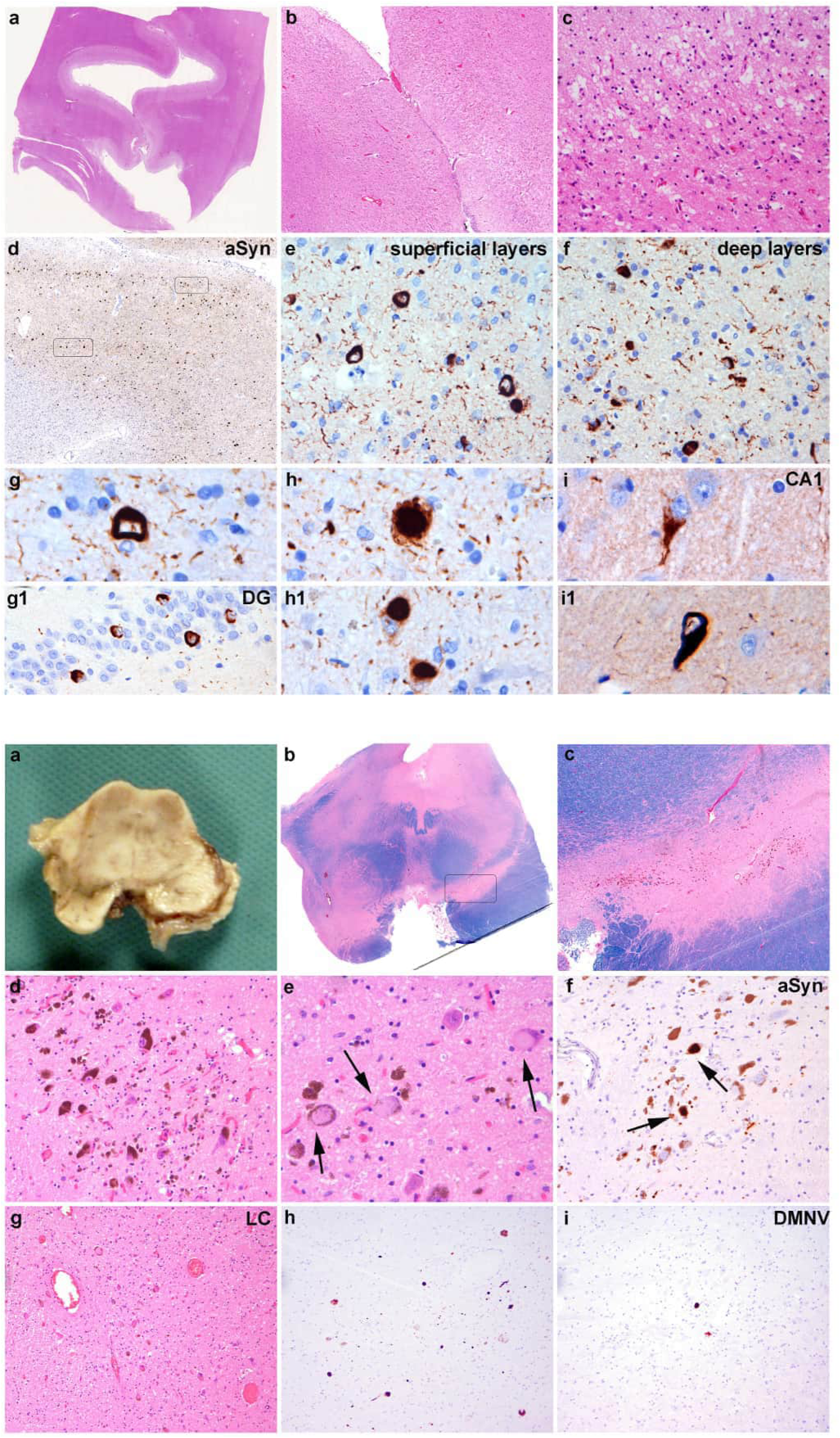
(Top panel) aSyn pathology in the cortical and hippocampal regions. A-C: HE- stained sections of the cingulate cortex show thinning of the cortical ribbon (A), neuronal loss preferentially involving the upper third of the cortex (B), and laminar superficial spongiosis (C). D-I: Immunohistochemistry for aSyn reveals a high pathology density with a spectrum of morphologies of the inclusions: ring-like with abundant fine neurites in superficial cortical areas (E), half-moon shaped in deeper layers (F), again ring-like in the dentate gyrus of the hippocampus (G, g), alternating with more compact and spherical (H, h) or tangle-like in pyramidal neurons of the CA1 sector of the hippocampus (I,i). (Bottom panel**) aSyn pathology in the midbrain.** A: Horizontal section through the midbrain reveals moderate pallor of the s. nigra. B, C, E: HE-stained sections show a moderate loss of pigmented cells of the s. nigra and locus coreuleus with extracellular pigment (B), and some cytoplasmic pale bodies (C, arrows) displacing neuromelanin granules. Interestingly, immunohistochemistry for aSyn (D, F) shows only mild pathology in the form of some diffuse and spherical cytoplasmic inclusions in the S.N. (C, arrows) and a few more in the L.C., associated with some neurites (F).

No co-pathology associated Aβ42-amyloid or TDP43 aggregates were detected. There was a mild tau-positive pathology in temporo-medial regions with some neuropil threads, few neurofibrillary tangles and pre-tangles, as well as isolated ballooned neurons in the amygdala and isolated oligodendroglial coiled bodies in the periamygdalar white matter, without grain pathology or astrocytic pathology.

### G14R aSyn mutation induces local structural alterations

To investigate how the G14R mutation affects structural properties of aSyn, we first performed in silico analysis using prediction models.^30–32^ According to the predictions, the G14R substitution reduces the percentage of α-helixes, increases β-strand and coil content, and augments the probability of β-sheet aggregation in the residues that follow immediately after (Figure S2). Experimentally, we performed NMR analysis using recombinantly produced G14R aSyn and compared it to recombinant WT aSyn. The two-dimensional NMR 1H/15N-correlation spectra showed narrow signal dispersion for both proteins, indicating their intrinsically disordered nature (Figure 2A). Although most cross peaks aligned between the two proteins, deviations were observed for cross peaks associated with residues near the mutation site (Figure 2, B-C). Upon sequence-specific analysis, it was clear that the perturbations in the NMR signal were confined to the region around the G14R mutation site as expected for an intrinsically disordered protein (IDP) (Figure 2C). Residues 14-20 are the mostly affected ones, which suggests charge-charge interactions between the positively charged side chain of G14R and the negatively charged side chain of E20 (Figure 2D).

**Figure 2.**
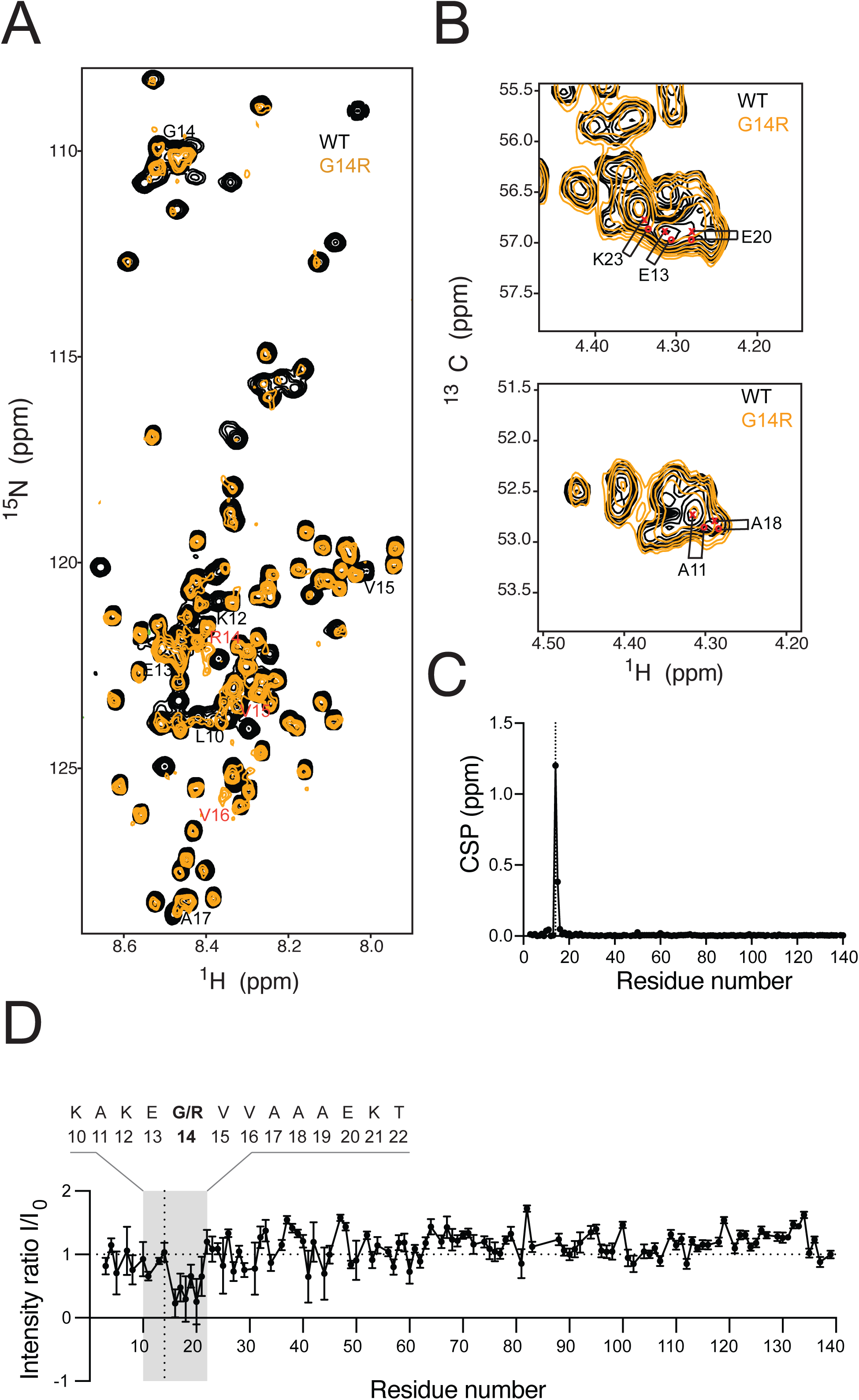
Effect of G14R mutation on aSyn structure. (A) 1H/15N-HSQC of wild type (WT, black) and G14R mutant (orange) aSyn. The affected residues are labeled. (B) Selected region of the 1H/13C HSQC of WT (black) and G14R (orange) aSyn. The most perturbed residues are labeled. (C) N-HN Chemical Shift Perturbations between WT and G14R aSyn based on the spectrum in a. (D) Residue-specific 1H/15N-HSQC peak intensity ratios for WT and G14R aSyn.

### Effect of the G14R mutation on aSyn aggregation in vitro and in cells

To assess the impact of the G14R mutation on aSyn aggregation, we first compared the fibrillization kinetics of both WT and G14R recombinant aSyn using an in vitro ThT-based aggregation assay. Interestingly, although the G14R showed a slightly faster aggregation rate, the aggregation curve for WT aSyn exhibited a higher ThT fluorescence suggesting potential differences in the nature or structure of the aggregates formed by these two variants (Figure 3, A-C). To extend the in vitro studies of aSyn aggregation kinetics, we investigated the aggregation propensity of the G14R mutation in cellular systems. For this purpose, we used the SynT/Sph1 model, an established system frequently employed to investigate aSyn aggregation (Figure 3D).^33^ Following the co-transfection of WT or G14R SynT variants with Sph1, human neuroglioma cells (H4) were immunostained to assess inclusion formation 48 hours post-transfection. Interestingly, the percentage of cells with aSyn inclusions increased significantly (>90%) in cells expression the G14R mutation when compared to WT aSyn (Figure 3E). Furthermore, small inclusions were predominantly present in the case of G14R with a higher inclusion number per cell. In contrast, a higher percentage of cells without inclusions were observed in cells expressing WT aSyn, and the area of inclusions was higher compared with G14R mutation (Figure 3, F-G).

**Figure 3.**
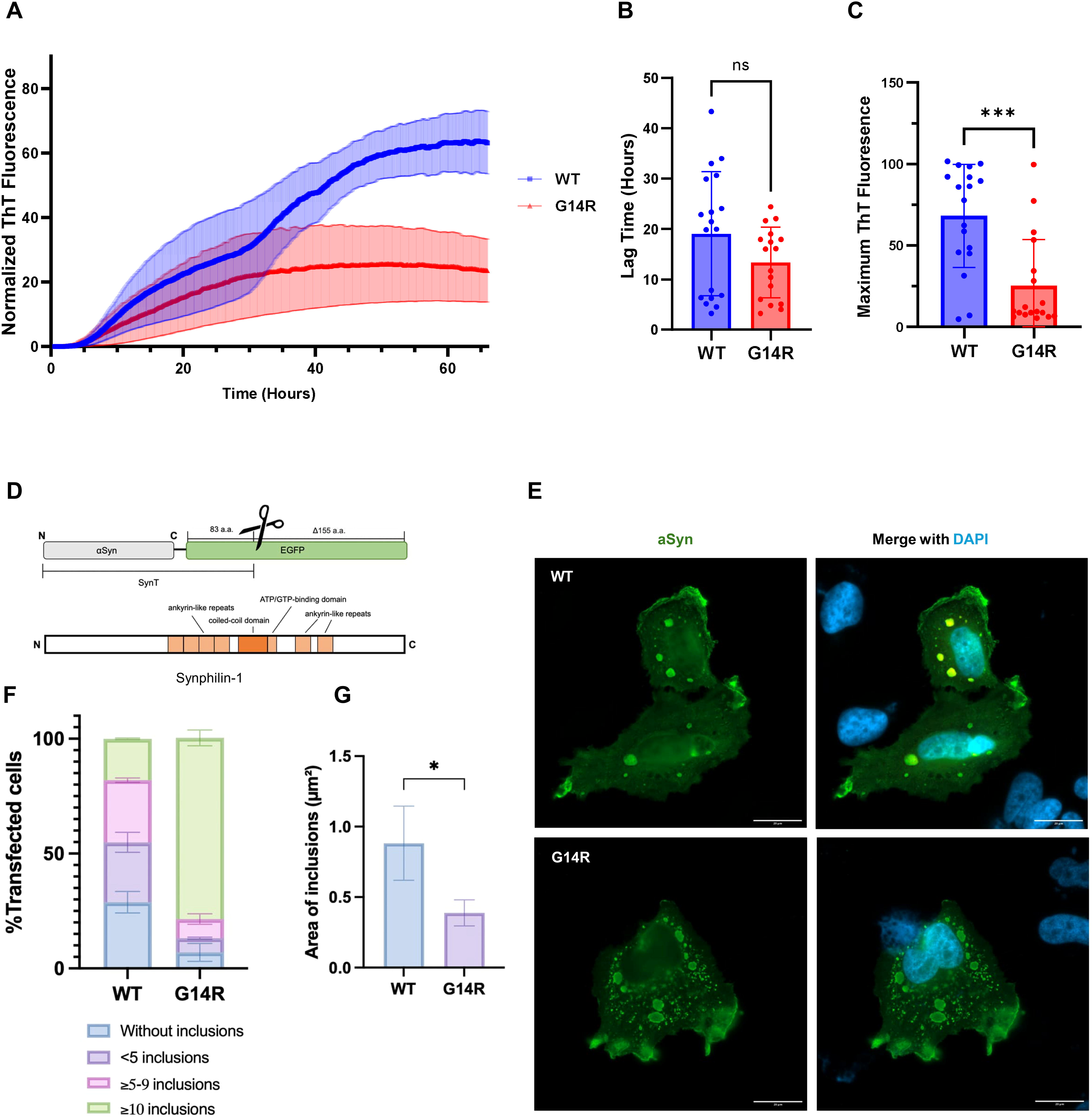
G14R aggregation propensity. A-C: ThT-based aggregation assays. (A) ThT-based Aggregation kinetic curves of WT and G14R aSyn. Curves were normalized to the maximum fluorescence intensity for each run. (B) The lag time in hours for WT and G14R aSyn. Each dot represents an individual technical replicate of the total replicates from 5 independent runs. (C) Normalized maximum ThT fluorescence for WT and G14R aSyn. Each dot represents an individual technical replicate of the total replicates from 5 independent runs. Data are represented as mean±SEM (N=5). Comparisons between WT and G14R aSyn in B and C were done using student t-test. D-G: Effect of G14R mutation on inclusion formation in cells. (D) the aggregation model constructs used in the study. The model is based on the co-expression of SynT and synphilin-1. (E) Representative immunohistochemistry images of the patterns of inclusion formation in H4 cells for WT and G14R aSyn. Scale bar: 20 µm. (F) Quantification of the number of inclusions and their area (G). 50 cells in 100x objective were counted for each experiment. The cells were classified into four different groups according to the pattern of inclusion. Data were analyzed using student t-test, and presented as mean ± SEM (*N=3*).

The difference in the aggregation profile of G14R aSyn prompted us to further analyze the morphology, organization, and structural properties of the fibrils prepared in vitro by cryo-EM. In cryo-EM micrographs, WT aSyn fibrils appeared well dispersed facilitating their structural analysis (Figure 4A). Two-dimensional (2D) class averaging showed the twisted morphology of these fibrils and revealed two populations of different width (Figure 4C, Figure S3B). Further 2D classification demonstrated that wide fibrils consisted of two protofilaments (2PF), while narrow fibrils contained only one (1PF; Figure S3C). Interestingly, subsequent 3D classification suggested a different fold in 1PF vs 2PF fibrils (Figure S3D). 3D reconstruction was successful only for the 2PF polymorph, converging to a map of 2·7 Å resolution (Figure S5A), which allowed building an atomic model (Figure 4E, G, H). This revealed a fold previously observed in recombinant WT aSyn fibrils,^34,35^ where the interface between the protofilaments spans residues 45 to 59 and is stabilized by a salt bridge between lysine 45 (K45) and glutamic acid 57 (E57; Figure 4H).

**Figure 4.**
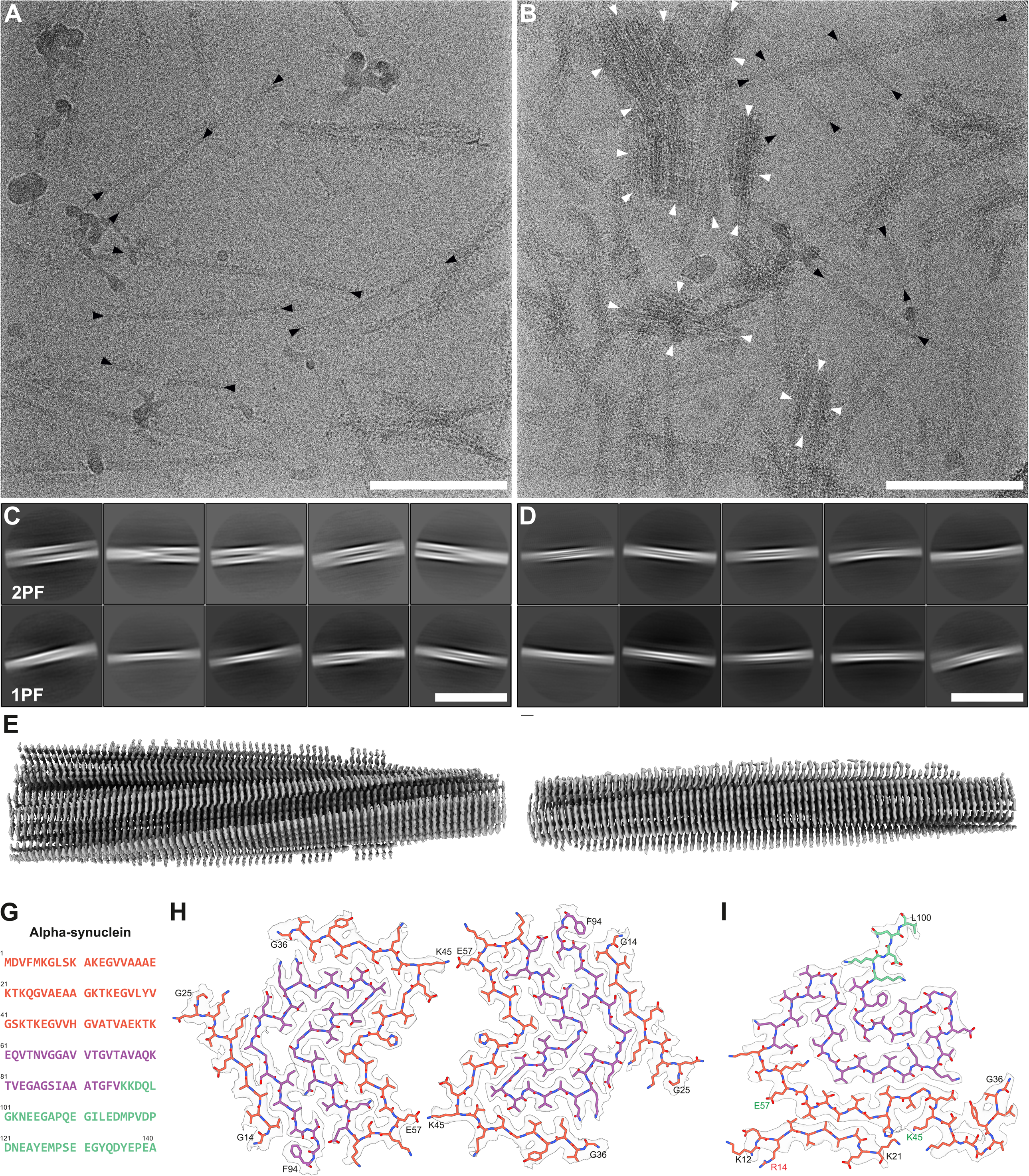
Characteristics of WT and G14R aSyn filaments. (A) TEM micrograph of wild-type (WT) aSyn amyloid filaments. Black arrows mark select filament ends. Scale bar: 100 nm. (B) TEM micrograph of G14R aSyn amyloid filaments. In the micrograph multiple aggregates consisting of laterally associated filaments can be seen. Black arrows indicate filament ends of exemplary filaments that were used for SPA processing. Scale bar: 100 nm. (C) 2D class averages (706 Å box size) of twisting WT aSyn fibrils, showing an interaction between two protofilaments. (D) 2D class averages (706 Å box size) of twisting WT aSyn filaments, showing interaction between two protofilaments (2PF) or a single protofilament (1PF). Scale bar: 50 nm. (E) Overview of the electron density map of WT aSyn filaments. (F) Overview of the electron density map of G14R aSyn filaments. (G) Amino acid sequence of human aSyn with distinct regions color-coded (N-Terminus in orange, middle hydrophobic region in purple, and C-Terminus in green). Scale bar: 50 nm. (H) The electron density map together with the atomic model of WT aSyn amyloid filaments featuring a single beta-sheet layer formed by two interacting protofilaments. The protofilament interface is stabilized by a K45-E57 salt bridge. (I) The electron density map together with the atomic model of a single beta-sheet of G14R aSyn amyloid filaments. The mutated residue is indicated in red, while residues forming the salt-bridge in the WT are marked in green.

In contrast to WT fibrils, most G14R aSyn fibrils associate laterally forming dense groups (Figure 4B, white arrowheads), limiting structural analysis to isolated fibrils (Figure 6B, black arrowheads). 2D classification revealed that essentially all G14R fibrils were formed by a single protofilament (Figure 4D; Figure S4B). Upon 3D classification and reconstruction to 3 Å resolution (Figure S5B; Figure S4 C, D), an atomic model was built (Figure 4F, I). The model was reminiscent of a different polymorph previously observed in WT aSyn fibrils,^35,36^ fibrils amplified from MSA seeds,^37^ and fibrils obtained from the brain of juvenile onset synucleinopathy patient.^38^ In this fold, the amyloid core extends into the aSyn C-terminal domain, adopting a classical Greek key topology. Consistently with our NMR data, residues around G14R occupy a completely different position in mutant vs WT fibrils. In G14R fibrils, an N-terminal stretch encompassing residues 13 to 20 binds the K45-E57 interface, thereby preventing the formation of the 2PF fibrils observed in WT. Although 3D reconstruction of WT 1 PF fibrils was not successful (see above), comparison of 3D classes between G14R (Figure S 4D) and WT 1 PF fibrils (Figure S3D) revealed their similarity. Altogether, these data suggest that while WT aSyn is highly polymorphic resulting in at least two stable conformations (1PF, 2PF), the G14R mutation strongly favors the 1PF fold, possibly by blocking the K45-E57 protofilament interface by an N-terminal stretch.

**Figure 5.**
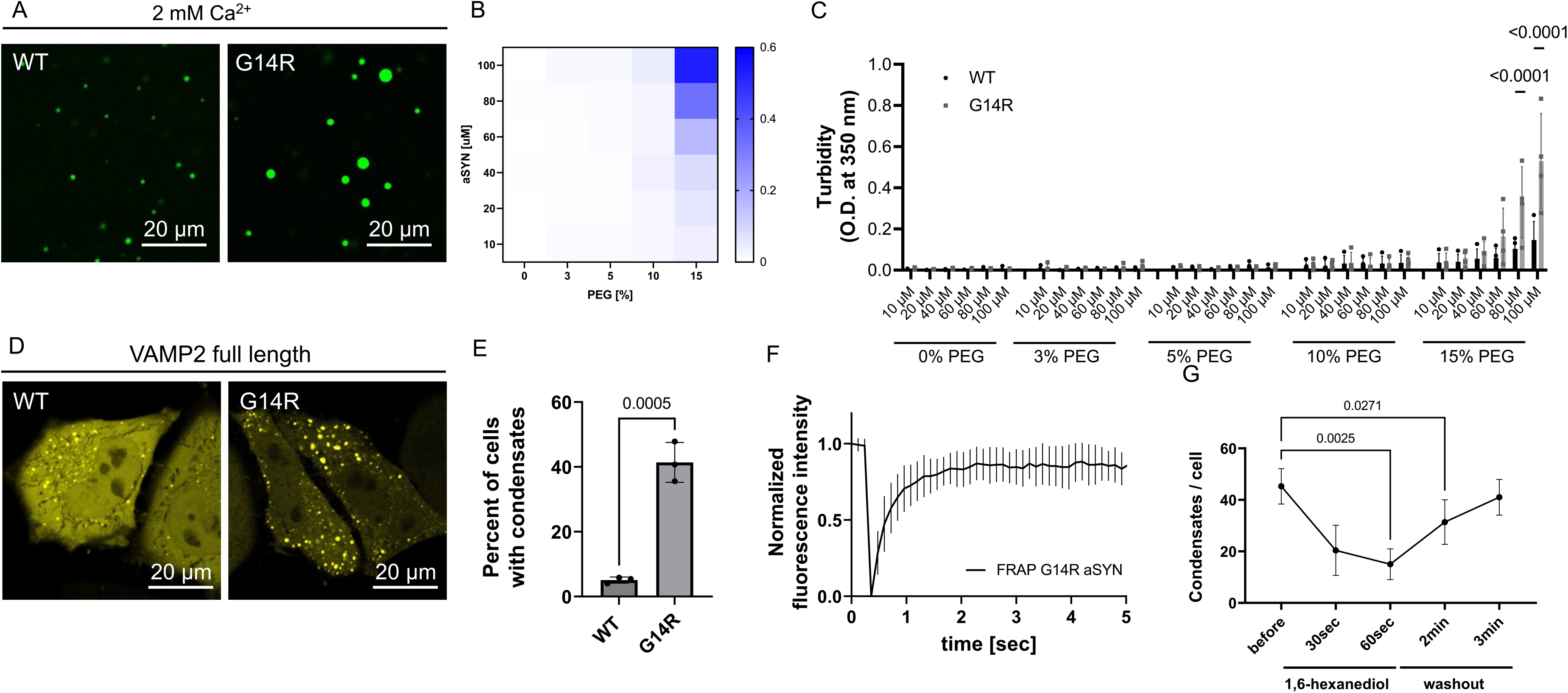
aSyn G14R shows increased condensate formation in vitro and in cells. (A) aSyn phase separation in the presence of 2 mM Ca^2+^ and crowding with 15% PEG 8000, immediately after PEG addition for aSyn wildtype (WT) and the disease variant aSyn G14R. aSyn concentration used: 100 µM. (B) Heatmap for turbidity measurements of aSyn phase separation in the presence of 2 mM Ca^2+^. Data derived from 4 independent repeats. (C) Comparison of aSyn phase separation derived from (B) showing increased condensate formation for the aSyn G14R disease variant. n=4, n represents independent repeats. Data are represented as mean +/- SEM. 2way ANOVA, Šídák’s multiple comparisons test. (D) Condensate formation of aSyn WT YFP and aSyn G14R YFP upon ectopic expression with VAMP2 in HeLa cells. aSyn G14R YFP shows increased condensate formation in cells. (E) Quantification of condensate formation. Data derived from incuCyte screening, 16 images per well, 3 wells per biological repeat, 3 biological repeats. n indicates biological repeats. Data are represented as mean +/- SD. Unpaired two-tailed t-test. (F) Quantification of fluorescence recovery after photobleaching (FRAP) of aSyn G14R YFP condensate in cells. Data are represented as mean +/- SEM. 3 biological repeats, n=11, n represents individual FRAP experiments. (G) aSyn G14R YFP condensates show dispersal and recovery upon incubation with 3% 1,6 hexanediol. n=8, n represents individual cells.

**Figure 6.**
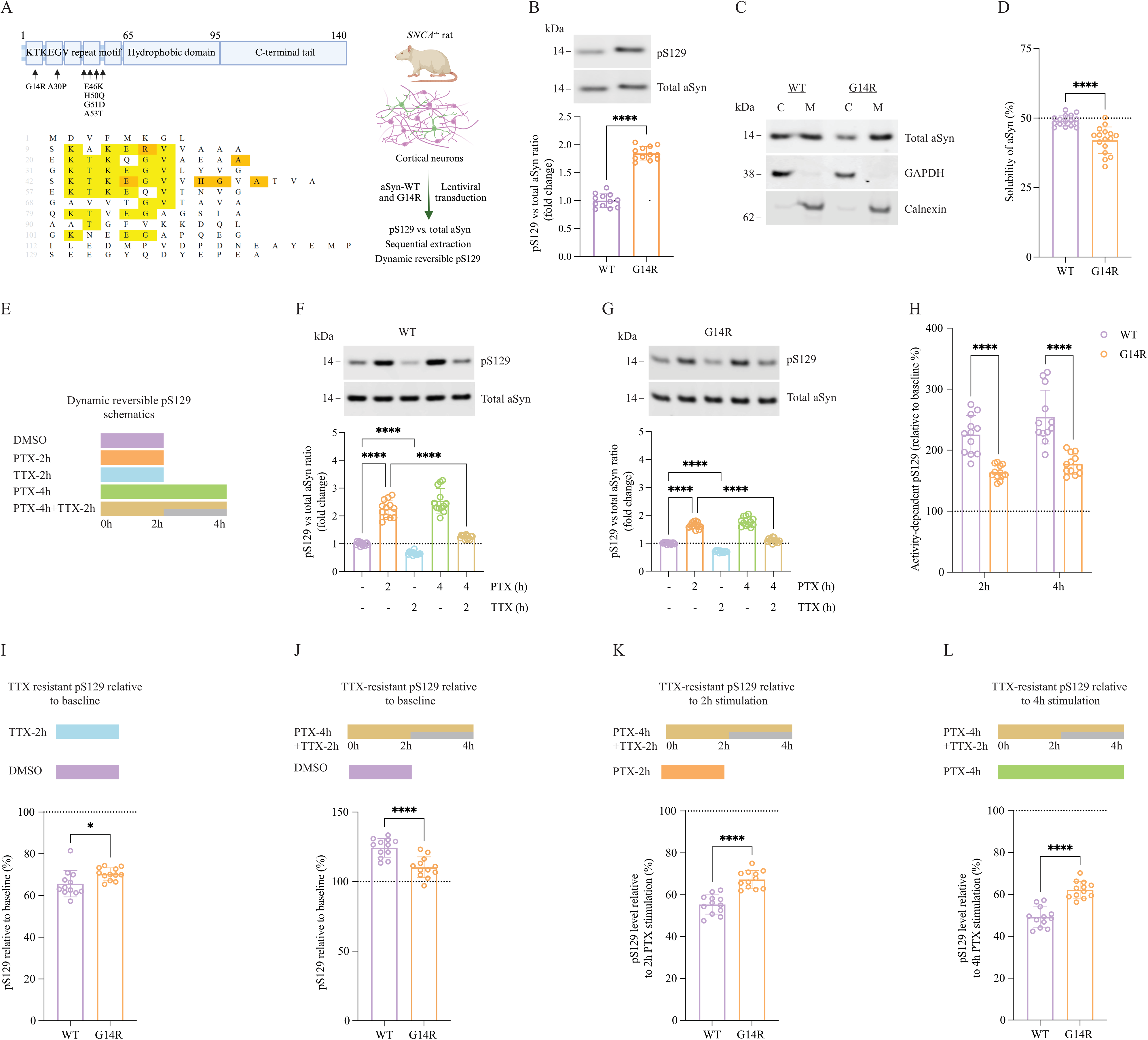
Dynamic activity-dependent pS129 of G14R and WT aSyn. (A) Schematic representation presenting the structure of aSyn including the KTKEGV repeat motif harboring most missense mutations associated with familial PD, the central hydrophobic region, and the C-terminal region (left top). Shown below is the alignment of aSyn sequence, where conserved amino acids of the KTKEGV motif are marked in yellow, and familial PD mutation sites are highlighted in orange. The experimental setup is displayed on the right. (B) Representative western blot (WB) for total aSyn and pS129 from DIV17-21 rat *SNCA*^−/−^ cortical neurons that express WT and G14R aSyn by lentiviral transduction. The quantification analysis from WB is shown below. (C) WB of WT and G14R transduced rat *SNCA*^−/−^ cortical neurons that at DIV17-21 underwent on-plate sequential extraction to separate the cytosolic (C) and membrane (M) fractions. Total aSyn was detected using the MJFR1 antibody, and the controls for the cytosolic and membrane fractions were GAPDH and Calnexin, respectively. (D) quantification of the solubility of WT and G14R aSyn from WB presented in (C). (E) Overview of the experimental conditions to investigate the dynamic reversibility of pS129. Details are present in the main text. (F-G) Neuronal activity-induced reversible pS129 (illustrated in schematic E) was observed in DIV17-21 rat *SNCA−/−* cortical neurons transduced with WT and G14R aSyn, respectively, using 20 µM picrotoxin (PTX) for stimulation and 1 µM tetrodotoxin (TTX) for inhibition. WB for quantifying total aSyn and pS129 was employed. (H) The percentage of increase in pS129 relative to baseline for WT and G14R aSyn after 2 h or 4h PTX stimulation (derived from F to G). (I) The percentage of TTX-resistant pS129 in WT and G14R variants (derived from F to G) relative to the basal state (DMSO vehicle). (J) The percentage of irreversible pS129 relative (derived from F to G) relative to the basal state (DMSO vehicle). (K) The percentage of irreversible pS129 relative to 2 h PTX stimulation (derived from F to G). (L) The percentage of irreversible pS129 relative to 4 h PTX stimulation (derived from F to G). Three independent experiments were performed on different days, with a total of 10-12 biological replicates. Data presented in B, D, and I-L are statistically analyzed with an unpaired *t*-test with Welch’s correction, while data in F-G were analyzed with Brown-Forsythe and Welch ANOVA with Dunnett’s T3 *post hoc* test for multiple comparisons. Data in H were analyzed with 2way ANOVA with Šídák’s multiple comparisons test. \*\*\*\**P*[<[0.0001; \*\*\**P*[<[0.001; \*\**P*[<[0.005; \**P*[<[0.05; ns, not significant. The error bar was mean ± SD.

### aSyn G14R undergoes condensate formation in vitro

To assess the impact of the G14R mutation on aSyn condensate formation, we tested aSyn droplet formation in the presence of Ca^2+^ as described previously.^39^ We find that aSyn G14R undergoes increased condensate formation (Figure 5A). To quantitatively assess this, we performed turbidity measurements, again in the presence of Ca^2+^ and at PEG concentrations ranging from 0 to 15% and aSyn concentrations ranging from 10-100 µM (Figure 5B). Here, aSyn G14R shows increased phase separation when compared to aSyn WT (Figure 5C).

### aSyn G14R forms cellular biomolecular condensates upon co-expression with VAMP2

To test condensate formation in cells we ectopically expressed aSyn YFP and VAMP2 in HeLa cells as shown previously.^39^ Both aSyn WT and aSyn G14R show condensate formation (Figure 5D) and quantitative evaluation demonstrates increased condensate formation for the aSyn G14R variant (Figure 5E). While condensate formation was overall increased, we found that the condensates formed by aSyn G14R still retained high mobility as demonstrated by fluorescence recovery after photobleaching (FRAP) experiments (Figure 5F). We found about 73% recovery one second after photobleaching and about 86% recovery five seconds after photobleaching, which is congruent with our and other previous reports.^39,40^ Furthermore, when cells were subjected to 1,6-hexanediol, a small aliphatic alcohol,^41–44^ aSyn clusters showed dispersal which reassembled after brief washout periods (Figure 5G), demonstrating that the observed clusters are dynamic structures.

### Effects of the G14R mutation on aSyn S129 phosphorylation

Next, we assessed whether the G14R mutation impacted on aSyn phosphorylation at serine 129 (pS129), which may impact not only pathology but also the physiological function of aSyn.^25^ Primary rat *SNCA*−/− cortical neurons were lentiviral-vector transduced to express either WT or G14R aSyn (Figure 6A). The pS129 status was first measured under normal unstimulated conditions. Interestingly, the G14R mutant exhibited a pronounced increase in pS129 levels when compared with WT aSyn (Figure 6B). The efficient phosphorylation of aSyn on S129 was recently reported to be inversely proportional to its solubility status, and it was shown that familial PD-associated aSyn mutants with more cytosolic (C) localization showed reduced basal pS129 levels.^25^ Since we found an increase in pS129 for the G14R mutant, we next assessed whether this increase could be related to the accumulation of aSyn at cellular membranes (M). Consistent with this hypothesis, we found the G14R mutant to be enriched in membrane fractions (calnexin fraction, ∼ 60 %) (Figure 6C and D).

After assessing the basal levels of pS129, we investigated the dynamics of this phosphorylation. According to recent data, elevated phosphorylation of S129 occurs in response to neuronal stimulation and is followed by a restoration of pS129 levels to baseline upon termination of the stimulus.^25^ Importantly, this dynamic reversibility of pS129 is reduced in neurons expressing PD-relevant A30P and E46K aSyn mutants compared to WT aSyn.^45^ Therefore, we subjected the WT and G14R aSyn transduced cortical cells to neuronal stimulation, inhibition, or a combination of both (Figure 6E). For the stimulation of cortical neurons, we used the GABA_A_ receptor antagonist picrotoxin (PTX), while inhibition was induced using tetrodotoxin (TTX), a sodium channel blocker that prevents action potentials. In line with our recent investigations for endogenous WT aSyn, the activation of neuronal cells resulted in a significant increase in pS129 at both 2 and 4-hour time points. Consistently, the inhibition of neuronal activity by TTX resulted in a decrease in the baseline levels by approximately 30%, and treatment with TTX 2 hours after PTX treatment was able to reverse the activity-induced elevation of pS129, restoring it to normal baseline levels (Figure 6F). Although we observed a similar pattern of activity-dependent pS129 for G14R aSyn (Figure 6G), the percentage of PTX-induced pS129 was higher in WT aSyn transduced neurons relative to basal levels (Figure 6H). To assess the dynamic reversibility of pS129, we compared 2h or 4h of PTX stimulation versus 4-hour PTX stimulation along with TTX inhibition applied halfway through PTX treatment. Interestingly, the percentage of irreversible pS129 levels in PTX/TTX-treated neurons was significantly higher for G14R relative to PTX stimulation for 2h or 4h (Figure 6, K and L). In other words, the reversal (dephosphorylation) of S129 phosphorylation is markedly compromised in the case of G14R compared to WT aSyn. Furthermore, G14R-transduced neurons were more resistant to TTX inhibition under unstimulated conditions (Figure 6I). In conclusion, our data indicate that the dynamic reversibility of pS129 is impaired in the disease-associated G14R aSyn mutant.

### G14R increases membrane localization without altering cytotoxicity in yeast

To further assess the effect of the G14R mutation on aSyn membrane interactions, we took advantage of the budding yeast, a model that affords the possibility to correlate subcellular distribution with toxicity.^46^ In accord with the results in neuronal cell cultures, we found that G14R aSyn was preferentially localized at the plasma membrane in yeast cells, when compared to WT aSyn, which was found at the membrane and also in cytosolic inclusions (Figure S6 A). The difference in localization did not alter the toxicity, which was identical to that observed for WT aSyn (Figure S6 B).

## Discussion

In the current study, we describe a new heterozygous aSyn variant, G14R, and provide detailed insight into the molecular effects of the mutation on aSyn biology and pathobiology. Interestingly, the patient presented with a complex neuropathological profile with clinical and neuropathological features not following the typical patterns of PD/DLB or MSA pathologies. A definite antemortem clinical diagnosis for the patient in our study was challenging due to the atypical clinical phenotype. The patient exhibited initial symptoms suggesting neurological alterations other than parkinsonism. However, the patient subsequently developed severe bradykinesia that did not effectively respond to L-dopa. Subsequent diagnostic tests indicated reduced dopamine transporter activity, which further supported the clinical suspicion of a complex neurodegenerative disorder associated with frontal dysfunction and parkinsonism.

Genetic testing identified the presence of the G14R missense mutation in aSyn. The mutation was present in the patient and one parent, supporting the hereditary nature of the condition. Although one of the parents was carrying the mutation, examination tests did not show signs of parkinsonism, behavioral disturbance, cognitive decline or RBD, suggesting incomplete penetrance. Importantly, the identification of the G14R aSyn mutation associated with the current disease, and the absence of this variant in any public genomic database as well as the evolutionary conservation of the G14 residue support the potential pathogenicity of the mutation. Importantly, the predicated and molecular findings reported in this study further support a pathogenic effect for the mutation. To date, several missense mutations as well as multiplications in *SNCA* have been reported to cause familial PD. These mutations are associated with PD pathology, and some variants seem to alter PD onset and/or severity, producing different phenotypes. *SNCA* multiplications and the A53T mutation lead to a more severe PD course.^47–49^ The G14R variant we identified was associated with rapid disease progression. The patient was bedridden after 4 years and died of complications 2 years later. Furthermore, severe bradykinesia and altered cognitive function presented shortly in follow-up exams, and initial assessments showed the presence of atypical symptoms including a stuttering speech with palilalia, myoclonic jerks, and action tremor of the limbs. Atypical phenotypes have also been reported in some aSyn variants, including G51D, that present with pyramidal signs, and rapid and severe disease progression.^15^

The neuropathological findings normally seen in PD/DLB and in MSA include pronounced degeneration of the SN and other brainstem nuclei, and aSyn pathology in the brain stem, cerebral cortex, as well as cerebellum in the case of MSA.^50–52^ This pathology is typically represented in the form of LB and LN in PD and DLB, a predominantly but not exclusive neuronal aSyn pathology, or GCI in MSA, a predominantly glial pathology, but also with frequent aSyn neuronal involvement. aSyn pathology is also present in several cases carrying *SNCA* missense mutations.^53,54^ In our case, we found severe nigral degeneration that likely accounts for the reported parkinsonian symptoms. However, we observed aSyn pathology that deviates from the classical morphology and distribution patterns of “classic” sporadic synucleinopathies. First, we found widespread aSyn pathology in different brain regions, including both superficial and deep layers of the cortex and the striatum. Our case displayed *SNCA* pathology that resembled individuals with the G51D and A53E missense mutations.^20,53^Furthermore, the typical condensed LB morphology was absent in the SN and other brainstem nuclei like the locus coeruleus or dorsal motor nucleus of the vagal nerve, and was hardly observed in the cortical areas. Instead, cytoplasmic inclusions with different morphological features were dominating, including ring-like, and comma-shaped accumulations, and few tangle-like inclusions in the hippocampus, among other morphologies. Interestingly, we observed some aSyn-positive GCI in the midbrain reminiscent of those found in MSA. G51D and A53E mutations were also been reported to be associated with atypical synuclein pathologies with overlapping features of both PD and MSA.^15,20,55^

The neuropathological observations in the index case align with the unusual clinical presentation. Our patient presented with symptoms like myoclony, dystonia, speech disorder and apathy that are suggestive of, or often seen in clinical phenotypes of FTLD, including corticobasal syndrome (CBS), progressive non-fluent aphasia (PNFA), and behavioral variant frontotemporal dementia (bvFTD).^56–58^ Although the patient did not fulfill the full diagnostic criteria of any of these syndromes, the initial clinical phenotype was, to some degree, related to CBS. Macroscopic and microscopic examination revealed an FTLD-pattern. Proteinopathies that are commonly found in FTLD include tau inclusions (FTLD-Tau), TDP-43 inclusions (FTLD-TDP) or, rarely, inclusions of the FET protein family.^59^ Interestingly, the neuronal inclusions related to the neurodegenerative changes in our study exhibited predominantly aSyn pathology, with only few independent tau aggregates in the limbic system, that might be related to age or other early and mild limbic tauopathy, without TDP-43, FUS or Aß aggregates. The association of widespread aSyn cytoplasmic inclusions with neuronal loss suggests that aSyn pathology contributes to the atypical clinical phenotypes presenting in this case. It is noteworthy to mention that there are limited cases of FTLD that are associated with LB pathology. Interestingly, one study reported severe FTLD in certain cases of atypical MSA, and the study proposed a new category for FTLD (FTLD-aSyn).^60^ Moreover, a novel *SNCA* mutation (E83Q) was reported recently, and this mutation was associated with clinical and neuropathological overlap features of DLB and FTLD.^61^ Overall, our findings support these studies and show the relationship of the novel mutation with a broader spectrum of aSyn pathology than previously thought.

As an IDP, monomeric aSyn exists in a dynamic ensemble of conformations in solution, while cellular aSyn exhibits a dynamic equilibrium between its cytosolic soluble disordered form and helically structured membrane-bound conformations.^62,63^ Several PD-associated mutations result in structural alterations that affect the solubility status of the protein and, consequently, functional and pathological properties.^33,64^ Similar to most known aSyn mutations, this mutation occurs in the N-terminal domain of aSyn which contains a repeated KTKEGV consensus sequence that is a structural determinant and involved in the formation of amphipathic α-helical structure important for the binding to membranes. In our study, computational analyses indicated a large drop in the helical propensity of aSyn when glycine at position 14 is mutated to arginine, in agreement with previous studies that showed that the introduction of positively charged K (and possibly R) at the hydrophobic sites of KTKEGV repeats disrupt the formation of helices and alter membrane binding. ^65^ Experimentally, NMR studies revealed that in monomeric, soluble aSyn, the effect of G14R mutation on aSyn is locally confined around the mutation site, as expected for an intrinsically disordered protein. However, it is noteworthy to mention that the perturbations extended beyond the immediate proximity of the mutation site. One of the affected residues is glutamic acid (E) at position 20 suggesting the presence of charge-charge interaction between R14 and E20. Changes in the conformational dynamics induced by this mutation, as observed here, are expected to affect the functional properties of the protein including lipid binding and aggregation properties.

The aggregation of aSyn into insoluble amyloid fibrils is related to the pathogenesis of different synucleinopathies, albeit in ways that are not fully understood. Furthermore, different neuropathological presentations in mice can be caused by distinct strains of aSyn fibrils.^22^

Aggregation prediction algorithms predict decreased aggregation propensity and increased solubility for the G14R mutant aSyn. An additional positive charge is expected to increase G14R solubility and arginine has been used in several studies to suppress aggregation,^66–70^ Interestingly, WT aSyn displayed a higher ThT aggregation profile although the lag time was slightly lower for the G14R. The observed differences align with the prediction algorithms and suggest differences in the structure of fibrils between the two variants, and the kinetics of fibrilization. Similarly, lower aggregation propensities have been reported in PD-associated mutation G51D.^27^ Although G14R and G51D occur on different sites, they are both present in the hydrophobic half of the helix, and glycine is substituted with a charged residue.

The cryo-EM study confirmed differences in the morphology and structural arrangements of fibril strains formed. With G14R, the fibrils tend to associate laterally, and most fibrils appeared to be composed of single protofilaments. In contrast, lateral association of WT fibrils was minimal, and two protofilament fibrils were also abundant. Similar polymorphs have been described.^34–38^ Furthermore, mutants that interfere with the formation of salt bridges showed a decreased tendency to form fibrils compared with WT aSyn.^71^ In addition, it has been reported that different aSyn strains or polymorphs can be associated with different neuropathologies, and the characterization of these strains can help in the diagnosis of different synucleinopathies.^36,38,72^ Thus, the G14R mutation likely leads to conformational changes that disrupt the formation of stable salt bridges between β-strands of aSyn, promoting the lateral association of filaments, which suggests different assembly mechanisms. In the cell-based system used, the G14R aSyn formed a larger number of inclusions, albeit of reduced size, when compared to WT aSyn. This possibly reflects the reduced lag phase observed with recombinant aSyn, and the reduced ThT binding of the fibers. In in vitro assays, aSyn G14R shows increased condensate formation, in line with increased condensate formation in cells upon aSyn and VAMP2 co-expression. This is consistent with what is known for the G51D variant, which has been associated also with atypical clinical and neuropathological phenotypes, exhibiting reduced in vitro aggregation but increased inclusion formation in cells.^33^ These findings highlight the importance of using different model systems for assessing the overall profile of PD-associated mutations, as some effects are likely to be context-dependent.

pS129 has long been widely recognized as a marker of pathology due to its presence in the vast majority of aggregated aSyn within LBs.^24^ Recently, two studies uncovered a physiological role of S129 phosphorylation.^25,73^ The phosphorylation of S129 under physiological conditions is a neuronal activity-dependent dynamic process.^25^ In other words, pS129 levels increase in response to neuronal stimulation, and the dephosphorylation of S129 back to baseline levels occurs rapidly once the neuronal activity is over. According to recent studies, the relevance of this process may be the dynamic reversibility rather than the phosphorylation itself. In our study, we found that G14R follows activity-dependent phosphorylation similar to the WT aSyn. However, two main differences exist compared to WT aSyn. First, the activity-dependent phosphorylation of S129 was less pronounced in the case of G14R. Second, pS129 dephosphorylation was impaired with the G14R variant. Collectively, we found that the dynamic reversibility of aSyn S129 is impaired for this novel disease-associated mutant. Our results are consistent with recent findings on PD-known mutations E46K and A30P, suggesting the dynamics of pS129 might be a physiological process that is compromised under pathological conditions.^45^

In conclusion, we have identified a novel heterozygous *SNCA* mutation that is associated with complex and atypical clinical and pathological phenotypes, characterized by the presence of widespread neuronal loss and FTLD-associated aSyn pathology. The functional data showed G14R mutation alters aSyn structure, changes the aggregation propensity of aSyn, and impairs physiological pS129 reversibility. Collectively, this new mutation supports the hypothesis that aSyn pathology is broader than previously thought, and that clinical features of synucleinopathies are likely not limited to parkinsonism and dementia, but can overlap with features of other neurodegenerative disorders. The study also sheds new light into how normal aSyn physiological functions can be impacted in the presence of disease-associated mutations, and opens novel avenues for understanding the molecular mechanisms associated with synucleinopathies.

## Abbreviations

aSyn: Alpha-synuclein
FTLD: Frontotemporal lobar degeneration LBD Lewy body dementia
MSA: Multiple system atrophy
PD: Parkinsońs disease
RBD: Rapid eye movement sleep behavior disorder
*SNCA*: Alpha-synuclein gene

## Data Availability

All data produced in the present study are available upon reasonable request to the authors

## Acknowledgements

We want to thank Dr. E. Friedrich for supporting and helping organizing the autopsy of our patient and Prof. T. Brücke for the initial care and referral of the patient. We are grateful to T. Cheng for assistance in cryo-ET experiments and T. Shaikh for advice on cryo-ET data processing.

Christof Brücke^1,2*#^, Mohammed Al-Azzani^3*^, Nagendran Ramalingam^4^, Maria Ramón^3^, Rita L. Sousa^3^, Fiamma Buratti^12^, Michael Zech^5,6^, Kevin Sicking^7,8^, Leslie Amaral^3,9^, Ellen Gelpi^2,10^, Aswathy Chandran^11^, Aishwarya Agarwal^11^, Susana R. Chaves^9^, Claudio O. Fernández^12^, Ulf Dettmer^4^, Janin Lautenschläger^11^, Markus Zweckstetter^13,14^, Ruben Fernandez Busnadiego^7,8,15,16^, Alexander Zimprich^1,2^, and Tiago Fleming Outeiro^3,14,17,18#^

## Author contributions

TFO, CB, and AZ contributed to the study conception and design. CB, MAA, NR, MR, FB, RLS, MZ, KS, LA, EG, AC, AA contributed to the acquisition of data.

CB, MAA, NR, KS, EG, UD, JL, MZ, RFB, AZ, TFO analyzed and interpreted the data.

CB, MAA, and TFO were involved in writing the initial draft of the manuscript. All authors reviewed and revised the manuscript.

## Funding

C.B. and E.G. were supported by local funding. A.Z. acknowledges grant support by the Michael J. Fox Foundation (MJFF-023915).

T.F.O. is supported by the Deutsche Forschungsgemeinschaft (DFG, German Research Foundation) under Germany’s Excellence Strategy, EXC 2067/1-390729940, and by SFB1286 (B8). N.R. and U.D. are supported by the National Institutes of Health (grant numbers NS121826, NS099328, NS109209, NS122880, and NS133979). M. Zweckstetter is supported by the Michael J. Fox foundation through MJFF-022411. M.Zech acknowledges grant support by the European Joint Programme on Rare Diseases (EJP RD Joint Transnational Call 2022), and the German Federal Ministry of Education and Research (BMBF, Bonn, Germany), awarded to the project PreDYT (PREdictive biomarkers in DYsTonia, 01GM2302), by the Federal Ministry of Education and Research (BMBF) and the Free State of Bavaria under the Excellence Strategy of the Federal Government and the Länder, as well as by the Technical University of Munich – Institute for Advanced Study. MZ’s research is supported by a “Schlüsselprojekt” grant from the Else Kröner-Fresenius-Stiftung (2022_EKSE.185).

R.F.B. is supported by the Deutsche Forschungsgemeinschaft (DFG, German Research Foundation) under Germany’s Excellence Strategy, EXC 2067/1-390729940, and by SFB1286 (A12). Cryo-EM instrumentation at the University of Göttingen was jointly funded by the DFG Major Research Instrumentation program (448415290) and the Ministry of Science and Culture of the State of Lower Saxony. K.S. and R.F.-B. were funded by the joint efforts of The Michael J. Fox Foundation for Parkinson’s Research (MJFF) and the Aligning Science Across Parkinson’s (ASAP) initiative. MJFF administers the grant ASAP-000282 on behalf of ASAP and itself.

J.L. is supported by the Royal Society (Royal Society Dorothy Hodgkin Research Fellowship, DHF/R1/201228), the Addenbrooke’s Charitable Trust (Grant Award 900325), the Leverhulme Trust (Research Project Grant, RPG-2022-257), as well as a Career Support Fund from the University of Cambridge.

## Conflict of interests

The authors have no relevant conflicts of interest to disclose related to the article.

## Data availability

All data needed to evaluate the conclusions in the paper are present in the paper and/or the Supplementary Materials.

The micrographs used for the single-particle analysis (SPA) of alpha-synuclein fibrils are available in the EMPIAR database under accession code EMPIAR-12518.

The atomic model and cryo-EM density map for the wild-type (WT) alpha-synuclein fibril are deposited in the Protein Data Bank (PDB) and Electron Microscopy Data Bank (EMDB) under accession codes 9HGS and EMD-52165, respectively. The corresponding data for the G14R mutant alpha-synuclein fibril are available under accession codes 9HGR (PDB) and EMD-52166 (EMDB).

A comprehensive Key Resource Table, detailing datasets, software, and protocols is available via Zenodo at https://zenodo.org/records/14734406. Additionally, the entry includes an Excel file with tabular data on the protofilament distribution of WT and G14R alpha-synuclein filaments, XML files containing tabular data for the FSC graphs of each density map used to determine the final resolution (generated with RELION 4.0), and two Python scripts for graphing the FSC XML data.

The full single-particle analysis protocol describing the cryo-EM data processing strategy is available at Protocols.io (https://www.protocols.io/view/single-particle-analysis-of-synuclein-fibrils-81wgbxozylpk/v1).

## Notes

### Competing Interest Statement

The authors have declared no competing interest.

### Author Declarations

We evaluated the patient at the Department of Neurology of the Medical University of Vienna, Austria, in a study with approval of the ethics committee of the Medical University of Vienna (EK1844/2019), and with written informed consent to participate in the genetic research described in this study by all subjects involved.

### Summary of Updates

The manuscript has been updated to change the license type to CC BY

