## Supplementary Material for "A novel alpha-synuclein G14R missense variant is associated with atypical neuropathological features"

### SUPPLEMENTARY METHODS

#### Neuropathology

Unfixed samples of selected regions were snap-frozen and stored at -80°C. The brain was then immersed in buffered 4% formalin for two weeks and multiple brain areas including the olfactory bulbs, all cerebral lobes (frontal, temporal, parietal, occipital), limbic system (cingulum, amygdala, hippocampus anterior and posterior), basal ganglia and basal forebrain (caudate, putamen, globus pallidum, nucleus basalis Meynert, subthalamic nucleus), thalamus, midbrain, pons, medulla oblongata, cerebellar vermis and hemispheres, and dentate nucleus, were embedded in paraffin and processed for histology. A panel of primary antibodies were applied in selected brain regions and included anti-alpha-synuclein (clone 5G4, Roboscreen, Leipzig, Germany), anti-phospho-Synuclein (Ser129, clone P-α-synuclein #64, Wako Chemicals), anti-ßA4 (clone 6F/3D, DAKO, Glostrup, Denmark), anti-phospho Tau (clone AT8, Thermo Scientific, Rockford, IL, USA), anti-phospho TDP43 (clone 11-9, CosmoBio, Tokyo, Japan), ubiquitin (polyclonal, DAKO), p62 (clone 3/p62 lck ligand, BD transduction Laboratories, Franklin Lakes, NJ, USA). The DAKO EnVision© detection kit, peroxidase/DAB, rabbit/mouse (Dako, Glostrup, Denmark) was used for visualisation of antibody reactions.

#### In-silico analysis

The presence of the p.Gly14Arg variant in public databases was investigated (PD Variant Browser and gnomAD),[1] and prediction tools were used for estimating variant pathogenicity (Combined Annotation Dependent Depletion (CADD), Polymorphism Phenotyping v2 (PolyPhen-2), evolutionary model of variant effect (EVE) and AlphaMissense.[2–4]

#### Expression and purification of recombinant WT and G14R aSyn

The introduction of the G14R mutation in the bacterial plasmid encoding aSyn was performed through site-directed mutagenesis (QuikChange II, Agilent) and verified by Sanger sequencing. Recombinant proteins were expressed in BL21(DE3) *E. coli* cells transformed with pET21A or pET28a vectors encoding either WT or G14R aSyn, as previously described.[5] The purification of the proteins included two chromatographic steps of anion-exchange and size exclusion chromatography (SEC) for aggregation assays and nuclear magnetic resonance (NMR) experiments. Purified aSyn was finally concentrated in SEC buffer (PBS, pH 7·4), sterile filtered, and stored at -80ºC. For NMR experiments, proteins were purified using 100 mM NaCl, 50 mM HEPES, pH 7·4, instead of PBS. The concentration of purified aSyn was determined by measuring the absorbance at 280 nm, employing the extinction coefficient of 5,960 M-1/cm-1 for aSyn.

#### Nuclear magnetic resonance (NMR) spectroscopy

NMR experiments were measured on a Bruker 700 MHz spectrometer equipped with a 5 mm triple-resonance, pulsed-field z-gradient cryoprobe using two-dimensional 1H,15N heteronuclear single quantum coherence (HSQC),[6] as well as 1H,13C heteronuclear single quantum coherence (HSQC) pulse sequences for monomer characterization at 15 °C. All experiments were performed in HEPES buffer (50 mM HEPES, 100 mM NaCl, pH 7·4, 0·02% $NaN_3$) with 10 % (v/v) D2O. The sample concentration for natural abundance experiments was 280 μM for both WT and G14R aSyn. Spectra were processed with TopSpin 3.6.1 (Bruker) and analyzed using Sparky 3.13 (T. D. Goddard and D. G. Kneller, SPARKY 3, University of California, San Francisco). The combined 1H/15N chemical shift perturbation was calculated according to (((δH)2+ (δN/10)2)/2)1/2.

#### In vitro thioflavin T fluorescence-based aggregation assays

Thioflavin T (ThT) based aggregation assays were performed in a CLARIOstar Plus plate reader (BMG Labtech; Ortenberg, Germany) using Costar black and clear bottom 96-well half area plates. Before starting the aggregation assay, a 500 μL master mix solution in PBS consisting of 50 μM monomeric WT or G14R aSyn, and 25 μM ThT was prepared, and 100 μL were pipetted into each well for a total of 3-4 replicates per condition. The aggregation protocol also included the previous addition of one 1-mm diameter glass bead to each well, and the use of PBS as a blank. The plate was sealed with microplate tape before transferring to the plate reader. The plate reader settings for aggregation were as follows: orbital shaking (60 seconds ON, 30 seconds OFF) for the plate with a frequency of 600 rpm at 37 °C and 5 minutes cycle (1000 cycles in total); ThT fluorescence intensity was monitored once per cycle using bottom optics with an excitation wavelength at 450 ± 10 and emission at 480 ± 10 nm. The aggregation curves were blank-corrected and normalized to the maximum fluorescence value for each run.

**Cryo-EM imaging and analysis of WT and G14R fibrils**

Fibril preparation

WT and G14R fibrils were prepared according to established protocols.[7] In summary, lyophilized WT and G14R aSyn were reconstituted in PBS to get a final concentration of 5 mg/ml. The protein samples were sterile-filtered, transferred into low-binding tubes, and incubated at 37 °C with a shaking frequency of 1000 rpm. After seven days, the tubes were collected and aliquoted under aseptic conditions.

Sample vitrification

Copper 200 mesh R2/1 EM grids (Quantifoil) were plasma cleaned (Harrick Plasma PDC-32G-2) for 45 s on medium power and mounted on a Vitrobot Mark IV (Thermo Fisher Scientific). Fibril preparations were diluted to a concentration of 0·5 mg/ml in 30 mM Tris-HCl buffer, pH 7·5. 3 µL of this solution were applied to each grid. Blotting was carried out at 10 °C and 100% humidity using a blot force of 7 and a blot time of 5 s.

Data collection

Cryo-EM datasets were collected on a Titan Krios cryo-transmission electron microscope (Thermo Fisher Scientific) equipped with a Falcon4i detector (Thermo Fisher Scientific) operating in counting mode. A nominal magnification of 130,000x was used, resulting in a pixel size of 0·92 Å. 4,092 and 6,474 movies were collected for WT and G14R aSyn, respectively. The nominal defocus range for both datasets was set between -1·2 to -2·4 µm. A total dose of 40 $e^{-}/Å^{2}$ was applied using an exposure of 6·59 and 6·66 $e^{-}/Å^{2}$*s, for WT and G14R aSyn, respectively. A Selectris energy filter (Thermo Fisher Scientific) with a slit width of 15 eV was used during data collection.

Data processing

Raw EER movies were fractionated, aligned, and summed using MotionCor2,[8] with a dose per frame of 1 $e^{-}/Å^{2}$. Contrast transfer function (CTF) parameters were estimated using CTFFIND4.[9] To pick segments on the WT dataset, a crYOLO[10] model was used that was previously trained on aSyn fibrils. Given the bundled morphology of G14R fibrils, another crYOLO model was trained on that dataset. For this, fibrils from approximately 50 to 100 micrographs were manually picked in RELION 4,[11] and the coordinates were exported to train a picking model in crYOLO. This model was then used to automatically pick fibrils in all G14R micrographs. All further processing was carried out in RELION 4. The picked filament coordinates were imported and used to extract helical segments with an inter-box spacing of ~15 Å, corresponding to approximately 3 asymmetric units on the amyloid filament. First, segments were extracted with a box size of 768 pixels and binned 3 times, resulting in a final box size of 256 pixels. These particles were subjected to 2D classification. After removing picking artifacts such as carbon edges, the remaining particles were used to extract unbinned segments with a box size of 384 pixels and a pixel size of 0·92 Å/pixel. Another round of 2D classification was performed, and only classes clearly displaying beta-sheets were selected for further analysis. These classes were used for 3D classifications, where helical parameters were determined by systematically scanning potential values, as the crossover distance was not visible in the micrographs. Multiple 3D classifications were run with a fixed helical rise of 4·75 Å and a twist ranging from -0·5° to -1·7° in approximately 0·05° steps. A featureless cylinder served as initial model. Classes that exhibited distinct polypeptide chains were used to generate class-specific initial models, which were further refined through additional 3D classification to select particles that matched the models. Once a class with separated beta-sheets was obtained, 3D refinements were conducted with a sampling interval of 1.8° and a T-value of 30. This was followed by CTF refinement and a final 3D refinement to achieve the best possible resolution and model quality.

Model building

WT and G14R density maps were imported into Coot,[12] where reference structures were introduced. For WT, the reference structure used was PDB 6RT0, while for the G14R filaments, PDB 8BQW was employed. For G14R fibrils, the G14R mutation was manually introduced within Coot. The atomic positions of the reference structures were refined in Coot. During this process, any residues present in the reference structures but not allocable in our density maps due to lack of detail were removed. Finally, the atomic models were subjected to additional refinement using Phenix.[13]

**aSyn aggregation studies in cells**

The investigation of inclusion formation in cells, based on the SynT/Sph1 aggregation model, was conducted as previously described.[14] Briefly, human neuroglioma cells (H4) were seeded into 12-well plates and transfected with equal amounts of plasmid DNA encoding for SynT and synphilin-1 (Sph1) 24 hours post-plating, as previously described. 48 hours post-transfection, cells were analyzed for inclusion formation through immunocytochemistry. 50 cells were counted per condition and the pattern of inclusions observed was categorized into 4 groups: cells that show no inclusions, cells with less than 5 inclusions, cells with 5-9 inclusions, and cells with ≥10 inclusions. The findings were represented as the percentage of the total number of transfected cells. Data were analyzed and presented as the mean ± SEM from 3 independent experiments using unpaired Student t-test.

**Solubility and dynamic pS129 reversibility experiments**

Plasmids and lentivirus production: WT or G14R aSyn synthetic cDNA sequences were digested by SpeI/NotI restriction enzymes and ligated into the respective sites of pLVX-EF1a-IRES ZsGreen1 (TaKaRa), which drives transgene expression by the EF1a promoter. Lentiviral packaging was carried out in 293-T cells as described (Ramalingam and Dettmer, 2021). Briefly, 293-T cells were transfected with WT or G14R plasmids along with pMD2.G and psPAX2 (packaging plasmids: Addgene #12259 and #12260, respectively). Culture supernatant containing viral particles was further purified/concentrated by ultracentrifugation at 100,000g. The viral pellet was then resuspended in neurobasal medium supplemented with B-27 and Glutamax (Gibco). On an average we obtained about $2{\cdot}5\times10^6$ viral particles per µL. To study the effect of G14R mutation on the phosphorylation status of aSyn at S129, primary cortical neurons obtained from *SNCA* knockout (*SNCA*−/−) E18 pregnant rats were cultured on poly-d-lysine coated 24-well plates and lentivirally transduced at DIV5 to express human WT or G14R aSyn. Neurons were cultured as described.[15] To assess the solubility of aSyn, sequential protein extraction to isolate cytosol (C) vs. membrane (M) protein fractions was carried out using the on-plate extraction technique as previously described.[16] For dynamic pS129 reversibility experiments, at DIV17-21, the cultures were exposed to vehicle (DMSO), 20 µM picrotoxin (PTX), 1µM tetrodotoxin (TTX), or a PTX/TTX combination, followed by cell lysis and immunoblotting to measure the levels of total and pS129 aSyn as previously described.[15,16]

**Yeast experiments**

Yeast plasmids

First, we introduced the G14R mutation in plasmids containing the cDNA for aSyn via Site-directed mutagenesis using QuickChange II Site-Directed Mutagenesis Kit (Agilent Technologies, SC, USA), following the manufacturer's instructions. Mutagenesis was performed in the plasmid backbone p426GPD encoding the WT aSyn-GFP and the plasmid p426GPD-GFP was constructed by inserting the GFP coding sequence as a *Spe*I-*Xho*I digested PCR product. All constructs were confirmed by DNA sequencing.

Yeast cell culture conditions and spotting assays

The *Saccharomyces cerevisiae* yeast strain BY4741 (*MATa his3Δ1 leu2Δ0 met15Δ0 ura3Δ0)* was transformed with plasmids by standard lithium acetate method. All strains were grown overnight in Synthetic Dextrose medium lacking uracil (SD-URA) (Takara Bio, Japan) at 30°C 180 rpm. To evaluate cell growth on solid media, cultures were grown to mid-log phase and normalized to equal densities, serially diluted 10-fold starting with an $OD_{600nm}$ of 1 and spotted on SD-URA agar plates. After 3 days incubation at 30 °C the plates were photographed.

Yeast cell microscopy

Yeast cell images were acquired with an epifluorescence microscope Zeiss Axio Observer equipped with a 100x oil objective lens.

**aSyn condensate formation experiments**

aSyn labelling

Labelling of aSyn was performed in bicarbonate buffer (C3041, Sigma) at pH 8 using NHS-ester active fluorescent dye AlexaFluor 488 5-SDP ester (A30052, Invitrogen Thermo Fisher). Excess-free dye was removed by buffer exchange using PD10 desalting columns (IP-0107-Z050.0-001, emp BIOTECH, Generon). Labelled protein concentrations were estimated using the molar extinction coefficient ε494 nm = 72,000 M−1cm−1.

aSyn phase separation assays

All aSyn phase separation assays were performed in 25 mM HEPES, pH 7·4. Phase separation was induced by mixing aSyn and PEG 8000 (BP223, Fisher Bioreagent) in the presence of calcium (21108, Sigma) as indicated. Images for phase-separated samples were acquired on an LSM780 confocal microscope (Zeiss, Oberkochen, Germany) using a 63x oil immersion objective. Zen 2.3 (black edition) and Zen 2.6 (blue edition) were used for data collection and image export. Images were taken at the indicated aSyn concentration, where aSyn was supplemented with 1% Alexa 488 labelled aSyn. For turbidity measurements phase separation samples were set up as described above using indicated concentrations of aSyn and PEG 8000 in the presence of 2 mM calcium (21108, Sigma). The turbidity of the samples was measured at 350 nm, 25 °C using 96-well Greiner optical bottom plates on a CLARIOstar plate reader (BMG LABTECH, Ortenberg, Germany) under quiescent conditions. CLARIOStar 5.01 was used for data acquisition. A sample volume of 100 µL was used, and readings were taken within 5 minutes of sample preparation. Raw turbidity data are plotted with background subtraction using GraphPad Prism 9.3.1. Data were obtained from four independent repeats.

Plasmids used in cellular phase separation studies

Wild-type human full-length *SNCA* and VAMP2, encoding aSyn and VAMP2, were cloned from cDNA obtained from human neuroblastoma cells (SH-SY5Y) and inserted into the pEYFP-N1 and pMD2.G vector (Addgene #96808, #12259) with a C-terminal YFP and Flag-tag, respectively. aSyn G14R was generated using KLD substitution (M0554S, NEB, Ipswich, US). All sequences were verified by sequencing.

Cell culture and transfection

HeLa cells were obtained from the European Collection of Cell Cultures (ECACC 93021013) and grown in Dulbecco's modified Eagle's Medium (DMEM) high glucose (31966-021, Gibco) supplemented with 10% fetal bovine serum (FBS, F7524, Sigma) and 1% Penicillin/Streptomycin (P0781, Sigma). Cells were grown at 37 °C in a humidified incubator with 5% CO2. Cells were tested for mycoplasma contamination using MycoStripTM (IvivoGen, Toulouse, France). Cells were plated at 20,000 cells/well in 8-well ibidi dishes (80807, ibidi, Gräfelfing, Germany) for confocal imaging or in 48-well plates (Cellstar, 677 180, Greiner bio-one) for incuCyte experiments. Cells were transfected the following day using Fugene HD Transfection reagent according to the manufacturer's protocol (E2311, Promega). Briefly, per reaction 12·5 µL OptiMEM (31985-062, Gibco) were set up in 1·5 mL sterile Eppendorf tubes. A total of 250 ng of DNA and 0·75 µL of Fugene reagent were added and incubated for 15 min at room temperature. The transfection mix was added to the cells for 1 min and then topped up with 300 µL complete media. Cells were imaged the next day.

Confocal microscopy and IncuCyte

Live cell confocal imaging was performed on an LSM780 microscope (Zeiss, Oberkochen, Germany) using a 63x oil immersion objective. YFP fluorescence was excited with the 514 laser at 2% laser power. Zen 2.3 (black edition) and Zen 2.6 (blue edition) were used for data collection and image export. For fluorescence recovery after photobleaching (FRAP) experiments images were taken with the 63x oil immersion objective, 20x zoom, 128x128 pixel resolution, at an imaging speed of 60 ms/image. Three pre-bleach images were acquired before the ROI was bleached with 100 iterations at 100% laser power using the 514 laser. Fluorescence recovery was recorded for 100 cycles. FRAP analysis was performed in FIJI using the FRAP profiler v2 plugin (Hardin lab, https://worms.zoology.wisc.edu/research/4d/4d.html). 1,6-hexanediol (240117, Sigma, USA) was prepared as 6% stock solutions in complete DMEM media and was added to the cells in a 1:1 ratio after the first image was recorded. Cells were imaged after 1,6-hexanediol was added, then media was removed and replaced with fresh complete DMEM media, and the same cells were imaged again. The number of condensates per cell was analysed using FIJI.[17] For quantitative evaluation of condensate formation cells were imaged with the IncuCyte S3 (Essen BioScience, Newark, UK). Phase brightfield and green fluorescence images were taken using a 20x objective at a 4-hour interval at 200 ms exposure, condensate formation (% of cells showing condensate formation) was evaluated 16 hours after transfection. IncuCyte 2021A was used for data analysis. At least three biological repeats with three technical repeats each were analysed blinded to the investigator.

Immunocytochemistry

HeLa cells were plated on 8-well Ibidi dishes (80807, ibidi, Gräfelfing, Germany), transfected as above, and fixed the following day using 4% paraformaldehyde in phosphate-buffered saline (PBS), pH 7.4. Blocking and permeabilization were performed using 10% FBS, 1% BSA and 0.3% TritonX-100 in PBS for 1h. Cells were stained using an anti-pS129 antibody raised in mouse (p-syn/81A, 825701, Lot: B318449, BioLegend), used at 1:1000 in PBS containing 1% BSA, and incubated overnight at 4 °C. Following three washes with PBS, secondary antibody (Alexa Fluor 594, A11072, Invitrogen) diluted at 1:1000 in PBS with 1% BSA was added to the cells and incubated at room temperature for 1h. After three washes with PBS, cells were imaged on an LSM780 microscope (Zeiss, Oberkochen, Germany) using a 63x oil immersion objective. YFP fluorescence was excited with the 514 nm laser, pS129 staining was detected using the 561 nm laser.

Quantification and statistical analysis

Data analysis and statistical analysis was performed using Excel 2016 and GraphPad Prism 9.3.1. Statistical parameters are reported in the Figures and the corresponding Figure Legends. Exact p-values are shown. Data distribution was assumed to be normal but this was not formally tested. No statistical methods were used to predetermine sample sizes but our sample sizes are similar to those reported in previous publications.[18–20] Samples were randomly allocated into experimental groups. Data collection and analysis have been performed blinded when indicated. Data were included if the control (wild-type) showed appropriate condensate formation.

### SUPPLEMENTARY FIGURES

A

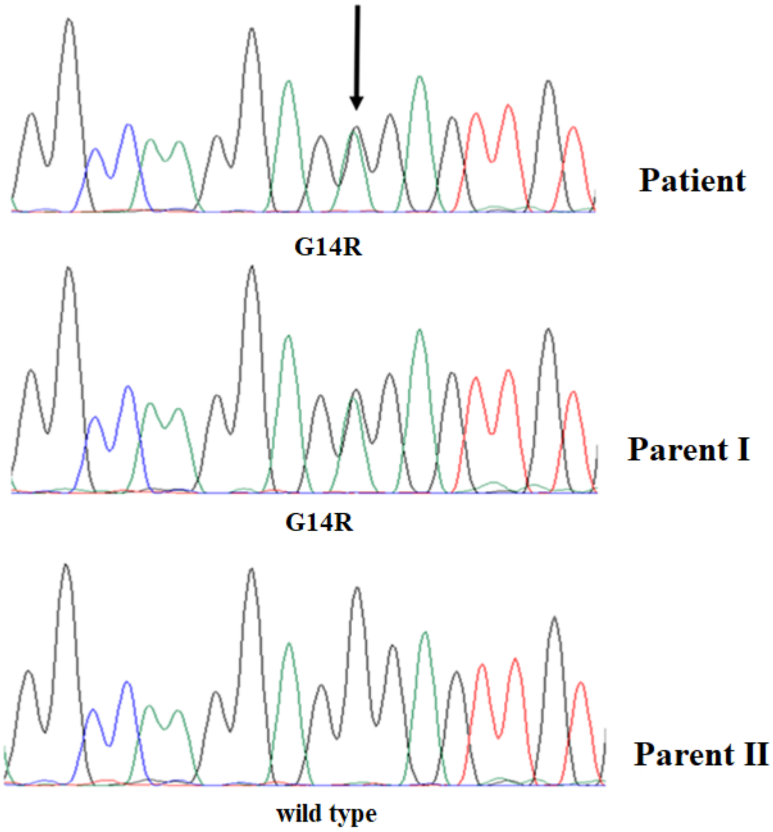

B

| | G14 | | | | | | | | | | | | | | | | |
|---|---|---|---|---|---|---|---|---|---|---|---|---|---|---|---|---|---|
| H. sapiens | K | G | L | S | K | A | K | E | G | V | V | A | A | A | E | K | T |
| P.Troglodytes | K | G | L | S | K | A | K | E | G | V | V | A | A | A | E | K | T |
| C.lupus | K | G | L | S | K | A | K | E | G | V | V | A | A | A | E | K | T |
| B. taurus | K | G | L | S | K | A | K | E | G | V | V | A | A | A | E | K | T |
| M.musculus | K | G | L | S | K | A | K | E | G | V | V | A | A | A | E | K | T |
| R.norvergicus | K | G | L | S | K | A | K | E | G | V | V | A | A | A | E | K | T |
| G.gallus | K | G | L | N | K | A | K | E | G | V | V | A | A | A | E | K | T |
| X.tropicalis | K | G | L | S | K | A | K | E | G | V | V | A | A | A | E | K | T |

**Figure S1. Identification of G14R mutation**. (A) Sanger sequence confirmation. The G14R was present in the patient and one parent; the other parent was found not to carry the mutation (Wild type). (B) Conservation of the SNCA G14R missense mutation among species. The NCBI Homolo Gene database (https://www.ncbi.nlm.nih.gov/homologene) was used to align protein homologues.

A

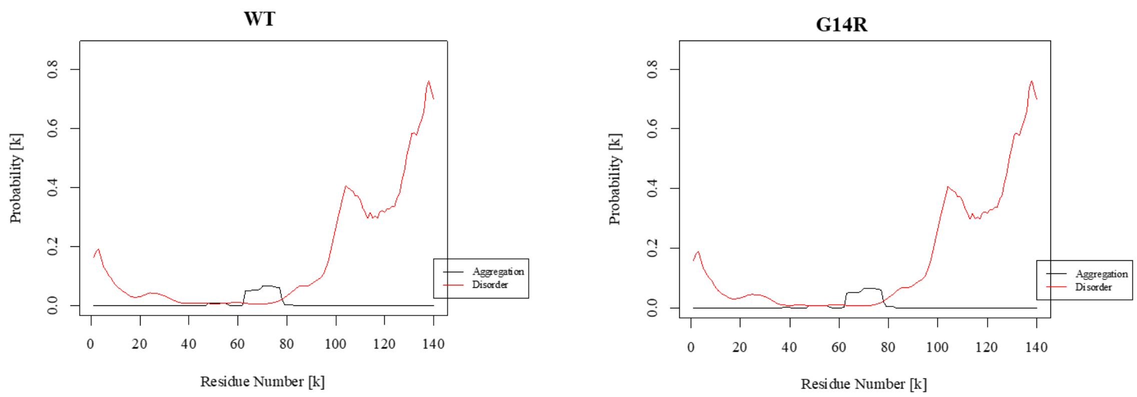

B

| Protein | best energy | % disorder | % α-helix | % β-strand | % coil |
|---|---|---|---|---|---|
| WT | -7.2393327 | 34.28 | 30.71 | 18.57 | 50.71 |
| G14R | -7.2393327 | 34.28 | 26.43 | 22.14 | 51.43 |

C

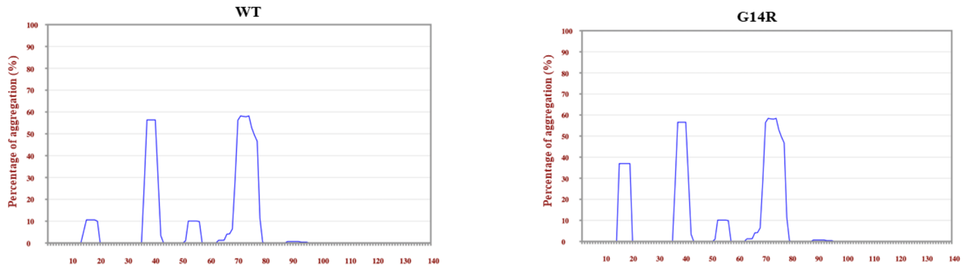

**Figure S2. Prediction for the effect of G14R mutation on aSyn structural properties**. (A) the effect of G14R mutation was assessed using PASTA 2.0 algorithm. G14R mutation is predicted to lead to a large drop in α-helix and an increase in β-strand and random coil as presented in (B). (C) G14R seems to cause an increase in the probability of β-sheet aggregation in the residues that follow immediately after according to the TANGO prediction algorithm.

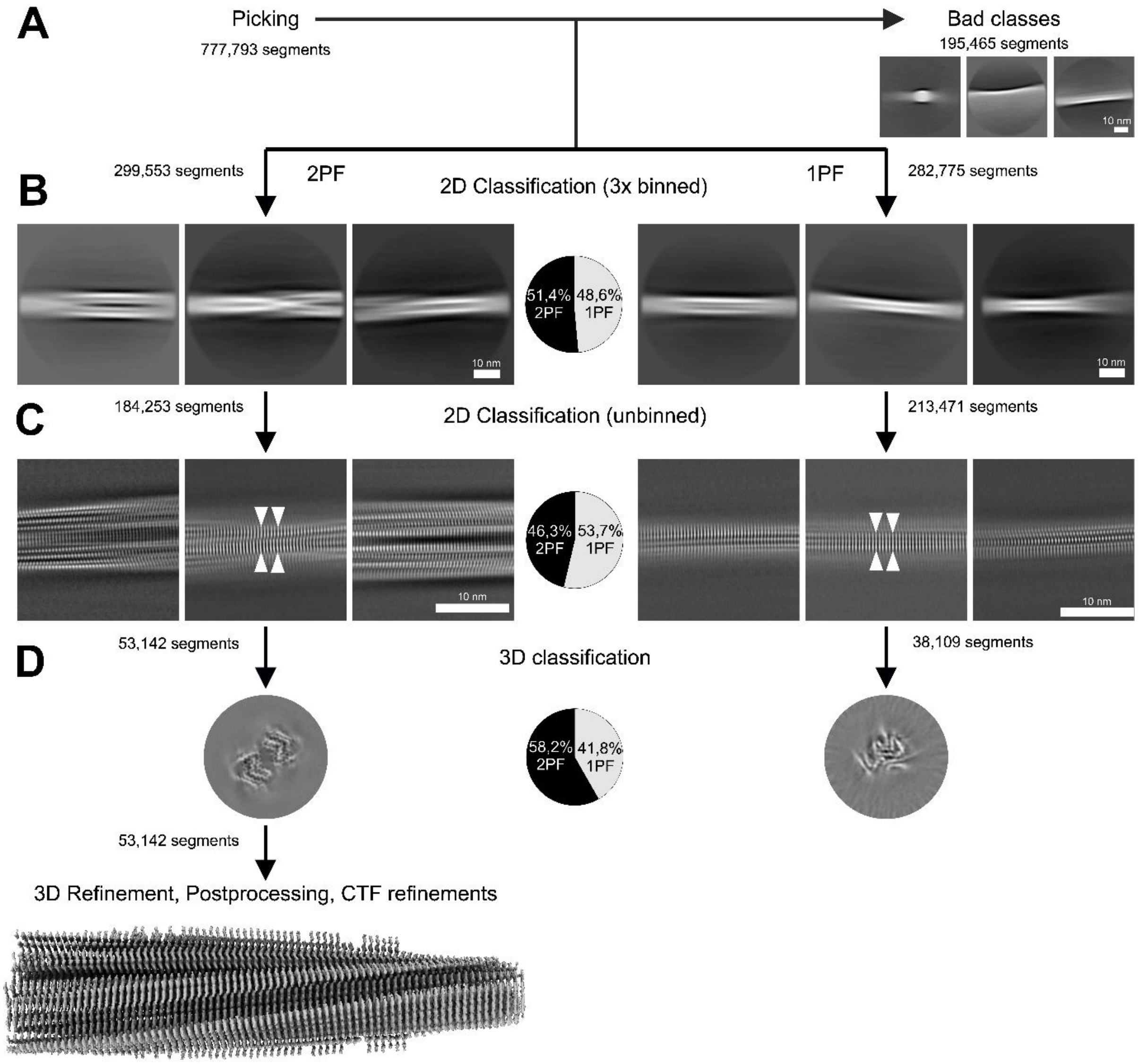

**Figure S3. Comprehensive Processing Workflow for the WT Dataset.** (A) Among the initially picked helical segments, a subset was discarded due to artifacts, such as segments containing carbon edges. The remaining segments were then categorized into two groups based on their structural characteristics: segments displaying two protofilaments (2PF, "wide") and those showing a single protofilament (1PF, "narrow"). (B) In the initial classification step using thrice-binned segments, the segments were almost evenly distributed between the two categories, as shown in the accompanying pie chart. (C) Subsequent classification using unbinned data corroborated the initial findings, confirming the even distribution of segments into the 2PF and 1PF groups, as also illustrated by the accompanying pie chart. (D) The results of the three-dimensional classification are presented, including the final electron density map for the 2PF data after 3D refinement, postprocessing, and CTF refinement. The accompanying pie chart indicates that a slightly higher number of segments contributed to the 2PF (two-protofilament) structure compared to the 1PF structure.

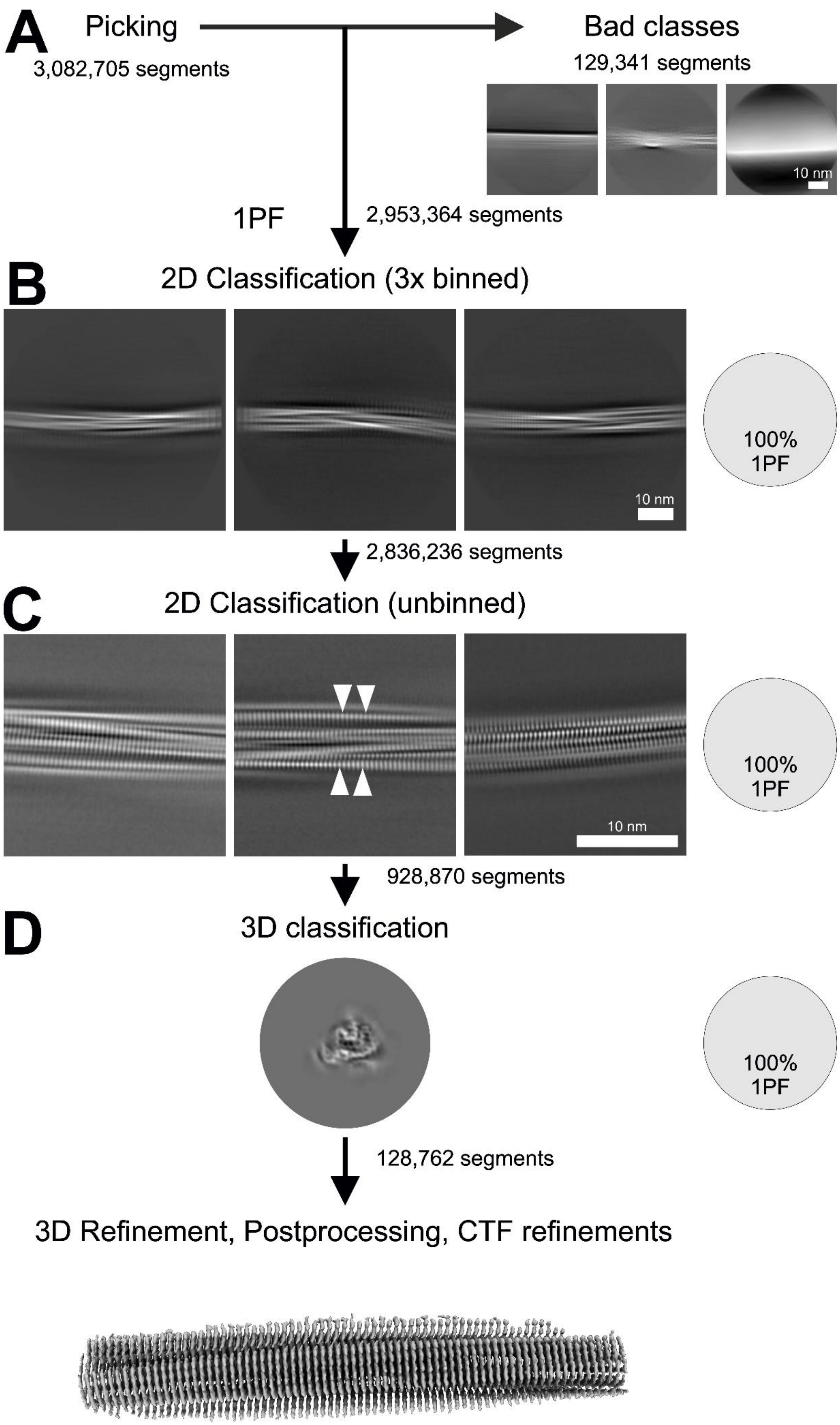
A
Picking
3,082,705 segments
Bad classes
129,341 segments
10 nm
1PF
2,953,364 segments
B
2D Classification (3x binned)
10 nm
100%
1PF
2,836,236 segments
C
2D Classification (unbinned)
10 nm
100%
1PF
928,870 segments
D
3D classification
100%
1PF
128,762 segments
3D Refinement, Postprocessing, CTF refinements

**Figure S4. Comprehensive Processing Workflow for the G14R Dataset.** (A) Among all the initially picked helical segments, a subset was discarded due to artifacts, such as segments containing carbon edges. The remaining segments were then categorized based on their structural characteristics, specifically those displaying a single protofilament (1PF, "narrow"). (B) In the initial classification step using three times binned segments, all segments were identified as consisting of a single protofilament, as shown in the accompanying pie chart. (C) Subsequent classification using unbinned data corroborated the initial findings, confirming that all segments are composed of a single protofilament. (D) Results of the three-dimensional classification, and the final electron density map after 3D refinement, postprocessing and CTF refinement.

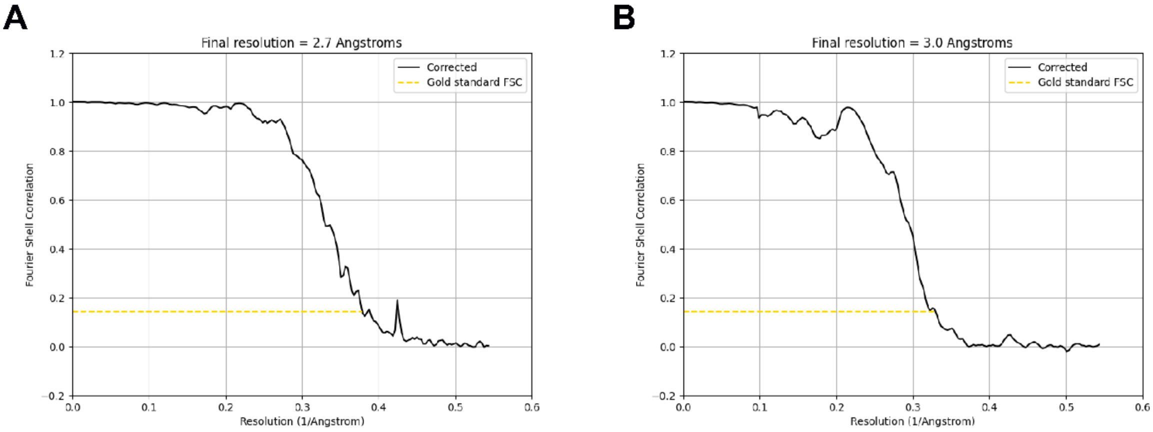

**Figure S5. Fourier Shell Correlation (FSC) curves for 2PF WT (A) and 1PF G14R (B).**

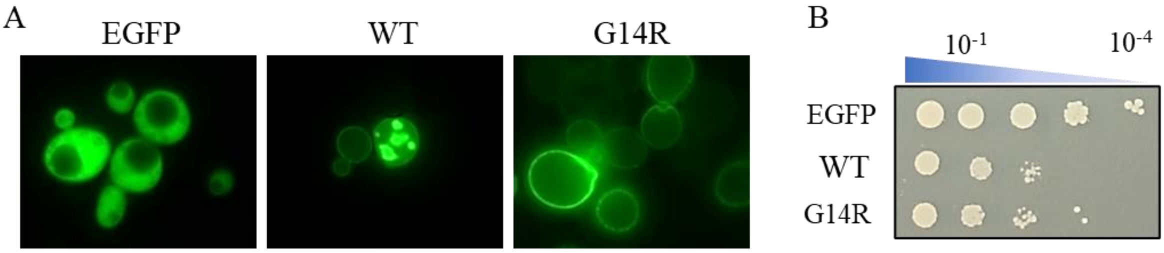

**Figure S6. Effect of G14R mutation on membrane interaction**. *S. cerevisiae* cells harbouring WT aSyn or G14R aSyn mutation were grown to the mid-log phase. (A) aSyn localization and inclusion formation were analyzed by fluorescence microscopy. (B) cellular growth was evaluated on solid SD-URA agar plates, where cultures were serially diluted 10-fold starting with an $OD_{600nm}$ of 1 and spotted on the plate.
